# Targeting mutant TNNT2-induced epigenetic perturbation and pathogenic signaling in left ventricular non-compaction cardiomyopathy

**DOI:** 10.1101/2024.10.09.24314670

**Authors:** You-Yi Li, Wei-Chieh Tseng, Hua-Ling Kao, Yu-Shin Shie, Sheunn-Nan Chiu, Ya-Ting Wu, Chung-Ming Sun, Shiou-Ru Tzeng, Liang-Chuan Lai, Miao-Hsia Lin, Yen-Wen Wu, Kuan-Yin Ko, Jyh-Ming Jimmy Juang, Ryan Hsieh, Mei-Hwan Wu, Wen-Pin Chen, Hong-Nerng Ho

**Affiliations:** Graduate and Institute of Pharmacology, College of Medicine, National Taiwan University; Taipei, Taiwan; Department of Pediatrics, National Taiwan University Hospital and Medical College, National Taiwan University; Taipei, Taiwan; Department of Applied Chemistry, National Yang Ming Chiao Tung University; Hsinchu City, Taiwan; Institute of Biochemistry and Molecular Biology, College of Medicine, National Taiwan University; Taipei, Taiwan; Graduate Institute of Physiology, College of Medicine, National Taiwan University; Taipei, Taiwan; Graduate Institute of Microbiology, College of Medicine, National Taiwan University; Taipei, Taiwan; Department of Nuclear Medicine, National Taiwan University Hospital and National Taiwan University College of Medicine; Taipei, Taiwan; Division of Cardiology, Department of Internal Medicine, National Taiwan University Hospital and Medical College, National Taiwan University; Taipei, Taiwan; Department of Mathematics, Durham University, Durham, United Kingdom; Department of Obstetrics and Gynecology, National Taiwan University Hospital and Medical College, National Taiwan University; Taipei, Taiwan; Taipei Medical University; Taipei, Taiwan

## Abstract

**Background:** Compound mutations of *TNNT2*^R141W/+^ (encoding troponin T) and *MYPN*^S1296T/+^ (encoding myopalladin) are associated with familial left ventricle non-compaction cardiomyopathy (LVNC). However, it remains unclear in which would be the pathogenic mutation, the underlying mechanism, and the target therapy for LVNC.

**Methods:** Knock-in C57BL/6J mice harboring mutations in orthologous genes in *Tnnt2*^R154W^ or/and *Mypn*^S1291T^ and human cardiomyocytes derived from iPSC of healthy donors and LVNC patients (LVNC-hCM) were employed for disease modeling, omics analysis, mechanistic study, and drug development.

**Results:** Using knock-in mice for disease modeling, it was clarified that the orthologous mutation in *Tnnt2*, but not in *Mypn*, led to cardiac hypertrabeculation, noncompaction, and heart failure. 3D protein structure modeling by Swiss-model found a loss of slat bridge between TNNT2(R141W) and E-257 in tropomyosin, contributing to the decreased cardiac contraction. Further mechanistic study discovered that troponin T (TNNT2) appears to function as an HDAC1 sponge in cardiomyocyte nuclei. The compromised association between nuclear TNNT2(R141W) and HDAC1 causes cardiac epigenetic perturbation and subsequentially leads to transcriptional dysregulation. The downregulation of cardiac muscular genes was concomitant with the impairment of cardiac contraction, which would be partially rescued by pan HDAC inhibitor. Besides, the upregulation of TGFβ-signaling molecules and EZH2 did contribute to cardiac growth defects, which were mitigated by TGFβR1 inhibitor (A83-01) and EZH2 inhibitor (GSK503), respectively. Simvastatin, a hit drug identified from the repurposed drug screening, can restore nuclear TNNT2(R141W)-HDAC1 association, thereby recovering cardiac epigenetic, translational profiles, growth and function in LVNC-hCM *in vitro* and cardiac function in LVNC mice harboring *Tnnt2*^R154W^ *in vivo*. The cardiac function was significantly improved in the proband receiving 5 mg once daily for consecutive two years.

**Conclusion:** Mutant TNNT2(R141W) diminished its nuclear HDAC1 sponge function in cardiomyocyte to induce LVNC pathogenesis through perturbating cardiac epigenetic and the gene expressions. Targeting to HDAC, TGFβ, EZH2 may rescue part of cardiac pathological signaling. Simvastatin can act as a chemical chaperone to comprehensively recover cardiac epigenetic via restoring nuclear TNNT2(R141W)-HDAC1 association.

## Introduction

Left ventricular non-compaction cardiomyopathy (LVNC) is characterized by a spongy left ventricular (LV) myocardium featuring abnormal trabeculations particularly in the left ventricular apex.^1, 2^ A population study found the mean annual incidence of newly diagnosed cases was 0.11 per 100,000 for children aged < 10 years and was highest in the first year of life (0.83 per 100,000 infants).^3^ The survival rate, unmarked by death or transplantation, stands at a mere 48% a decade following the diagnosis. Notably, patients who manifest symptoms during early infancy with a dominant dilated phenotype exhibit even graver long-term outcomes.^3^ Genetic mutations associated with LVNC encompass variations in MIB1^4^ and TBX20^5^, TAZ^6^, LMNA^7^, NUMB/NUMBL^8^, mitochondrial genome mutations (distal 22q11·2^9^), and the sarcomere-encoding genes (MYH7, ACTC1, TNNT2, MYBPC3, TPM1, LDB3, and TNNI3^10^). Nevertheless, the comprehensive mechanisms underlying the contribution of various genetic mutations to LVNC pathogenesis remain subject to further elucidation.

The dysregulation of Notch signaling, such as inactivation of cardiac Mib1, myocardial Jagged1, or endocardial Notch1^4^ along with the sustained upregulation of MFng and Dll4^11^, can precipitate premature trabeculation and myocardial non-compaction during the embryonic development of mice hearts. Conversely, the aberrant activation of TGF-β signaling, induced by mutant TBX20, has been observed to impede myocyte proliferation in mice embryonic myocardium, ultimately leading to LVNC^12^. Furthermore, cardiometabolic dysfunction had been identified as a constituent of the pathophysiological processes contributing to LVNC^13^. Nonetheless, the precise mechanism through which mutant sarcomere genes influence cardiomyocyte proliferation and myocardial compaction remain elusive.

In our investigations, we encountered a LVNC family simultaneously harboring mutations in TNNT2^R141W/+^ and MYPN^S1296T/+^. Despite the administration of leading-edge pharmaceutical interventions, including anti-congestive agents, diuretics, renin angiotensin blockade, beta-blockers, or mineralocorticoid receptor antagonists as well as antiplatelet or anticoagulation therapy to prevent thromboembolism^2, 14, 15^, the prognosis remained unfavorable. Mechanical circulatory support by devices, such as LV assist device, and cardiac transplantation, while considered as a therapeutic option for individuals resistant to pharmacotherapy^16^, frequently faces constraints stemming from limited donor resources.^1, 3^ Thus, we embarked on an integrated protocol that encompasses genetic characterization, disease modeling by knock-in mice harboring mutant genes, hiPSC modeling, omics study, and drug repurposing screening, with the aim of elucidating potential mechanisms-driven therapeutic strategies.

## Methods

### 1. Identify the index LVNC family with genetic information

LVNC patients who had advanced heart failure for more than 3 months during the infancy were enrolled (Figure 1A & supplementary Figure 1). Whole exon sequencing was performed to find out the genetic mutations, which were further confirmed by Sanger sequencing. From such a LVNC family, we identified two heterozygous missense mutations in TNNT2 (chr. 1:201333464 G>A; p.R141W) and MYPN (chr. 10: 68210378 A>T; p.S1296T) in the proband and her father.

**Figure 1.**
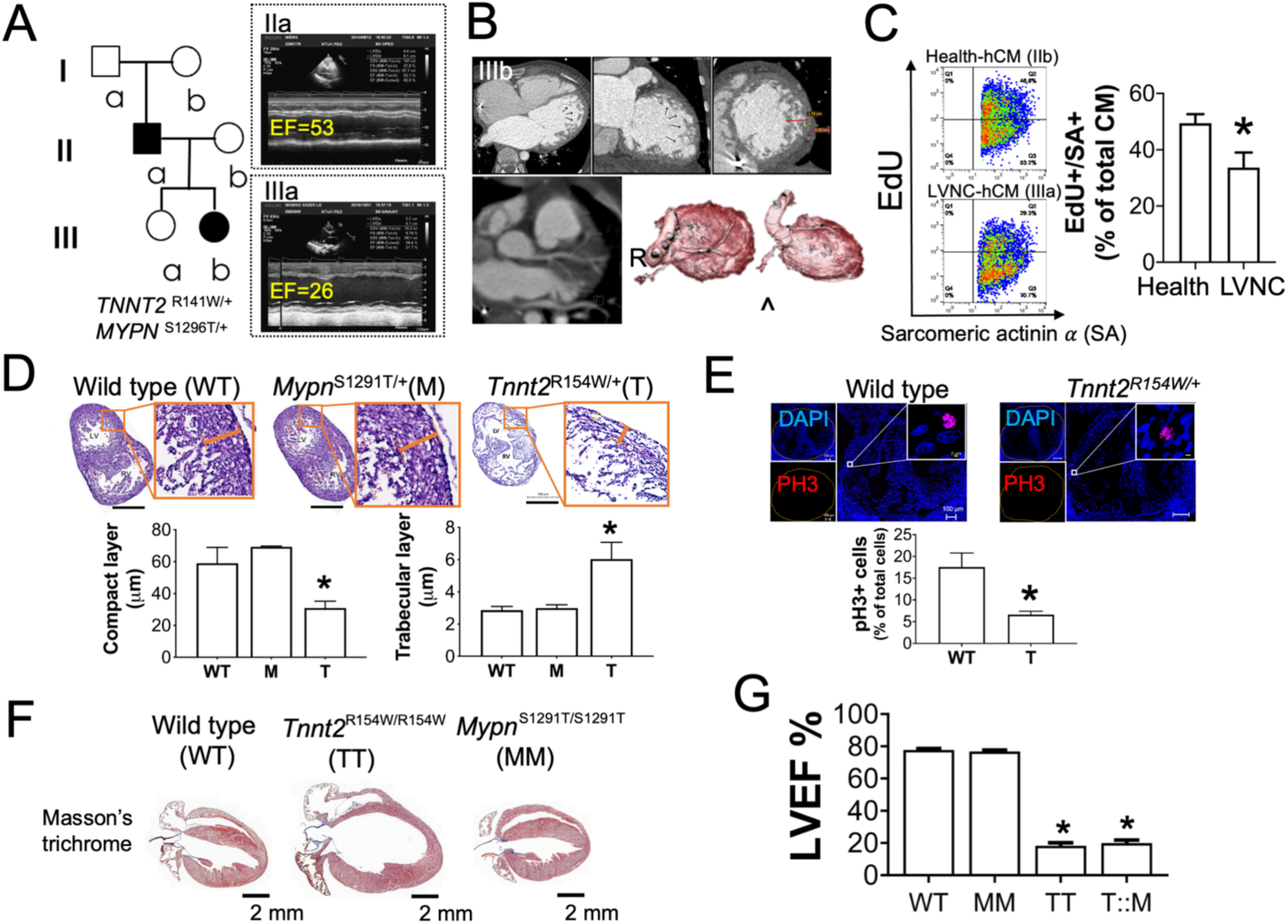
Characterization of disease phenotype in a LVNC family and mice harboring patients’ mutant genes. (A) Pedigree of LVNC family carrying *TNNT2*^R141W/+^ and *MYPN*^S1296T/+^ mutations. Representative acquired echocardiography images of LVNC patients (IIa & IIIb) in right panel were measured from the proband (IIIb, EF=26 %) and her father (IIa, EF=53.1 %). (B) The images of computed tomography (CT) reconstituted coronary arteries and the representative cardiac CT sections of the proband. The arrows indicate the representative LVNC features of hypertrabeculation and non-compaction. (C) The decrease of cell proliferation in human cardiomyocytes derived from the probands’ iPSC. Data are mean± SEM (n=10). *P<0.05, vs. health group by unpaired t test. (D) The decrease of the thickness in compacted myocardium and the increase of the length in intraventricular trabecula layer in E15.5 embryonic heart of *Tnnt2*^R154W/+^ (T) mice in comparison to those in wild type (WT) and Mypn^S1291T/+^ (M) mice. Data are mean± SEM (n=3). *P<0.05, vs. WT by one-way ANOVA with post-hoc Dunnett’s multiple comparison test. (E) A decrease of phospho-histone 3 (PH3) expression in myocytes of E15.5 mice carrying *Tnnt2*^R154W/+^ (n=8) as compared to those in WT group (n=9). Data are mean± SEM. *P<0.05, vs. WT group by unpaired t test. (F) Cardiac structure and fibrosis were examined by Masson’s trichrome stain in the hearts of WT and mutant (*Tnnt2*^R154W/R154W^ (TT) & *Mypn*^S1291T/S1291T^ (MM)) mice at the age of 8-9 weeks old. The scale bar represents 2 mm. (G) Comparison of left ventricular ejection fraction (LVEF) measured by echocardiography between WT and mutant groups of *Mypn*^S1291T/S1291T^ (MM), *Tnnt2*^R154W/R154W^ (TT), and *Mypn*^S1291T/+^::*Tnnt2*^R154W/+^ (T::M) at the age of 6 weeks old. Mice carrying *Tnnt2* ^R154W^ exhibited a significant reduction on LVEF. The data represent the mean± SEM (n=10 in each group). Statistical significance is denoted as *P<0.05, compared to WT, determined using one-way ANOVA with post-hoc Tukey’s test.

### 2. Human induced pluripotent stem cell (hiPSC) study

Human induced pluripotent stem cell (hiPSC) lines derived from LVNC patients (the proband: iH-LVNC-hiPSC, her father: iF-LVNC-hiPSC; the pedigree in Figure 1A & echocardiography in supplementary Figure 1), normal healthy woman (her mother: iM-hiPSC), and a normal healthy man (deidentified patient: health·m-hiPSC) were reprogrammed from peripheral blood monocytes (PBMCs) of the same patient by Sendai virus carrying four Yamanaka factors (CytoTune-iPSC 2.0 Sendai Reprogramming kit, Thermo Fisher Scientific Inc., MD, USA). All clinical investigations have been conducted according to the Declaration of Helsinki principles. All human studies have been approved by Research Ethics Committee D of the National Taiwan University Hospital. The proband’s informed consent was obtained from her parents. Chromosome analysis of hiPSC lines was performed by karyotyping (supplementary Figure 2). The hiPSC lines were cultured by monolayer in Matrigel-coated dishes and maintained in an Essential 8 culture medium (Thermo Fisher Scientific Inc., CA, USA) containing bFGF (10 ng/mL). Cardiomyogenic differentiation was conducted by the modified protocol of the chemically defined conditions^17^ based on the RPMI 1640 medium. In brief, CHIR99021 (6-12 μM) was placed in RPMI 1640 medium containing B27 Insulin minus supplement (RPMI/B27/I-, Thermo Fisher Scientific Inc., CA, USA) to induce mesoderm differentiation for two days. Then, the hiPSC differentiation medium was changed to RPMI/B27/I-containing IWP-4 (5 μM) to induce cardiomyogenic differentiation for the next two days. The differentiated cells were maintained in the RPMI/B27/I-medium until the presence of the beating myocytes around days 9-12 after differentiation. Cardiomyocytes derived from hiPSC (hiPSC-hCM) were further enriched by metabolic selection with glucose-free RPMI medium containing lactate (4 mM). The purity of cardiomyocytes was meticulously assessed through immunocytochemistry. Staining was conducted employing a mouse monoclonal antibody specific to sarcomeric actinin α (SA, Sigma-Aldrich), succeeded by the secondary antibody, Alexa Fluor 488-conjugated goat anti-mouse IgG. As a negative control, staining with Alexa Fluor 488 goat anti-mouse IgG alone was employed. Subsequently, the labeled cells underwent analysis using the BD LSRFortessa cell analyzer. For advanced experiments, only cell batches with a purity exceeding 90% of SA+ cardiomyocytes were employed. SA+ cardiomyocytes were subsequently reseeded onto Matrigel-coated dishes/plates and maintained in RPMI 1640 medium supplemented with B27 for the specified experiments.

### 3. LVNC mouse model and the cardiac function imaging

The homologous loci of human *TNNT2*^R141W^ and *MYPN*^S1296T^ were equal to murine *Tnnt2*^R154W^ and *Mypn*^S1251T^. Therefore, the orthologous *Tnnt2*^R154W/+^ and *Mypn*^S1291T/+^ knocked-in mice on C57BL/6j background were generated by Gene Knockout Mouse Core Laboratory of National Taiwan University Center of Genomic Medicine. Genetically homozygous *Tnnt2*^R154W/R154W^ (*Tnnt2*^R154W^) and *Mypn*^S1291T/S1291T^ (*Mypn*^S1291T^) were obtained from intrastrain crossing *Tnnt2*^R154W/+^ and *Mypn*^S1291T/+^ mice, respectively. Genetically heterozygous *Tnnt2*^R154W/+^::*Mypn*^S1291T/+^ LVNC mice were obtained from crossing *Tnnt2*^R154W/+^ and *Mypn*^S1291T/+^ mice. PCR and DNA sequencing were performed to confirm the genotype. All animal procedures and protocols followed the US National Institutes of Health and European Commission guidelines and were approved by Institutional Animal Care and Use Committee (IACUC) of National Taiwan University College of Medicine and College of Public Health. All animal experiments were conducted at an AAALAC-accredited facility.

Drug administration (Simvastatin: 8·27 μmol/kg/day; DMSO: 3·6 μL/day) was conducted by the implantation of an osmotic infusion pump (ALZET model 2006, Durect Corp. Cupertino, CA, USA) to the subcutaneous space of LVNC mice’s backs at the indicated age in the results. For continuous drug administration during day 1-126, the vehicle- or simvastatin-containing pump was replaced with a new one for every 42 days. Cardiac motion function of LVNC mice was measured by Prospect T1 echocardiography imaging system (Scintica Instrument, Inc., Webster, TX, USA) in M-mode under anesthetization by 1-2% isoflurane inhalation.

### 4. Immunocytochemistry and flow cytometric analysis (FACS)

EdU pulse-chase labeling was performed in hCM during day 12-16 after myogenic differentiation by Click-it Plus EdU conjugated with Alexa Fluor 594 or Alexa Fluor 647 flow cytometry assay kit (ThermoFisher Scientific, CA, USA). Cells were co-stained with mouse monoclonal antibody against sarcomeric actinin α (Sigma-Aldrich) in combination with rabbit polyclonal antibody against DLL4 (Genetex) or JAG1 (Abcam). The details of antibodies were listed in supplementary Table 1. The samples were followed by staining with secondary antibodies of Alexa Fluor 488 goat anti-mouse IgG and Alexa Fluor 594 donkey anti-rabbit IgG. The labeled cells were analyzed by BD LSRFortessa cell analyzer.

### 5. Pull-down assay and liquid chromatography-tandem mass spectrometry (LC-MS/MS)

The ORF of *TNNT2* coding either the wild type (WT) or missense mutation (R141W) was recombinated to multiple cloning site (MCS) of pDEST-flag-SBP-MCS (supplementary Figure 3). Plasmid of either pDEST-*TNNT2* or pDEST-*TNNT2*^R141W^ was transfected to healthy-hCM or LVNC-hCMs by Lipofectamine 3000 (Invitrogen, Life technologies) in Opti-MEM (GIBCO, Life Technologies) on day 14 after myogenic differentiation. The proteins associated with the fusion proteins of flag-SBP-*TNNT2* or -*TNNT2*^R141W^ were pulled down by Dynabeads® M-280 Streptavidin (Invitrogen, Life technologies). TNNT2-associated proteins were identified by LC-MS/MS (Orbitrap Fusion Lumos with ETD, Thermo Fisher Scientific Inc.) and were further validated by western blot or PLA assay.

### 6. Immunofluorescence (IF) or immunohistochemistry (IHC)

Mice E15.5 embryo or adult mice hearts were harvested from mice under anesthetization by 3-5 % isoflurane inhalation. After washing out blood, the embryo or heart was fixed by 4% paraformaldehyde on ice for 3h. Then, it was washed out by DPBS for three times. The samples were first incubated in 35 % sucrose until sedimentation, then embedded in OCT and stored at -80°C freezer before making cryosections with a thickness of 8 μm. After washing out OCT, the sections were further used for H&E stain, Masson’s trichrome stain or immunofluorescence stain followed by sequential staining with primary antibodies, including mouse monoclonal antibody against sarcomeric actinin α (Sigma-Aldrich), phospho-histone 3 (phospho-S10, Abcam), and the corresponding secondary antibody (Alexa Fluor 488 conjugated goat anti-mouse IgG & Alexa fluor 594 conjugated donkey anti-rabbit IgG). The details of antibodies were listed in supplementary Table 1. DAPI is used for nuclear counterstain. Fluoromount-G mounting medium (Invitrogen) was used to mount slides. The fluorescent images were acquired by LSM 880 confocal microscope system.

### 7. Immunoprecipitation (IP)

Mice ventricle tissue was cut into small pieces and homogenized in the homogenizer. Extracted cytosol and nuclei part lysate with the nuclear extraction kit (Abcam). Immunoprecipitation of Lamin B1 (1:250, GeneTex) or HDAC1 (1:250, GeneTex) using Pierce Crosslink Magnetic IP/Co-IP kit (ThermoFisher scientific) and immunoblot target antigen TNNT2 (1:20000, GeneTex) was subquentially performed.

### 8. Proximity ligation assay (PLA)

Mice hearts were fixed by 4% paraformaldehyde (PFA) on ice for 3h. After washing out PFA, mice hearts were immersed in 35 % sucrose until sedimentation at 4°C. Then the heart was embedded in OCT and was stored at -80 °C until the manufacture of frozen sections with the thickness of 8 μm. After OCT removal by PBS and antigen retrieval by HistoReveal (Abcam), cardiac sections were blocked by Donkey anti-mouse IgG (1:40 in PBS containing 0.1 % Triton X-100, overnight at 4 °C). After washing out, cardiac sections were incubated in PBS containing dual-stain primary antibody cocktail: (1) mouse monoclonal antibody against troponin T (1:500, GeneTex) and rabbit polyclonal antibody against HDAC1 (1:300, GeneTex), or (2) mouse monoclonal antibody against troponin T (1:500, GeneTex) and rabbit polyclonal antibody against Lamin B1 (1:100, GeneTex) at 4°C overnight. After the primary antibodies were washed out with PBS for three times, Duolink PLA Fluorescence kit (Sigma-Aldrich) was used to identify the colocalization of the proteins in interest listed above by following the manufacturer’s instructions. DAPI was used as the nuclear counterstain. High resolution of PLA image was acquired by LSM880 confocal microscope system in Airyscan mode with 3D Z-stack for the advanced analysis of the localization/distribution and the volume of PLA spot by Imaris Surfaces (Oxford Instruments plc, Tubney Woods, Abingdon, Oxon OX13 5QX, UK).

### 9. Measurement of myocyte contraction and intracellular calcium transient

Myocyte shortening was measured by IonOptix myocyte contractility system (IonOptix LLC., MA, USA) with electrical pacing at 1 Hz in the absence or presence of isoprenaline at the concentrations of 0.03, 0.3 and 1 μM and was shown as the percent shortening of the cell length. Intracellular calcium transients were recorded under optical pacing at 1 Hz by the high-content imaging system in hiPSC-hCM expressing GECIs (genetically encoded calcium indicator) and ChR2-K-GECO (channelrhodopsin-2) delivered by lentivirus and AAV, respectively^18^. Cells were kept in the system with 5% CO2 supplementation at 37 C for 2 h equilibrium before recording and were optically paced at 1 Hz.

### 10. Omics study

#### Proteome analysis

The total proteins of hiPSC-derived cardiomyocytes treated with or without simvastatin were extracted by RIPA buffer containing protease and phosphatase inhibitor cocktails (Thermo Fisher Scientific Inc., USA). The samples were further digested by trypsin. After the samples were desalted and dried, LC-MS/MS analysis was performed by LTQ-Orbitrap Velos mass spectrometer system. The peptide and protein group IDs were obtained by MASCOT database searching with precursor mass accuracy of 7 ppm and MS/MS accuracy of 0.5Da. The large mass-spectrometric raw data were further analyzed by MaxQuant software based on the Uniprot human database with label-free quantification method and followed by the statistical analysis using Perseus software.

#### Transcriptome analysis

The total RNAs of mice hearts or hiPSCs derived cardiomyocytes were extracted by TRIzol reagent (Invitrogen, Thermo Fisher Scientific Inc., USA) and were further purified from the upper aqueous layer of the TRIzol-chloroform homogenate by Quick-RNA kit (Zymol Research, CA, USA).

For RNA sequencing of mice ventricles, it was performed by NovaSeq 150PE platform, generating 20M reads/sample to characterize the transcriptional profile of mice hearts (wild type (WT) versus *Tnnt2*^R154W^ (TT) group, four mice in each group).

For RNA sequencing of hiPSCs derived cardiomyocytes treated with or without simvastatin, TruSeq Stranded mRNA sequencing was performed by Phalanx Biotech. In brief, polyA mRNA from an input of 500 ng high quality total RNA (RIN value > 8) was purified, fragmented, and the first- and second-strand cDNA were synthesized. Barcoded linkers were ligated to generate indexed libraries. The libraries were quantified using the Promega QuantiFluor dsDNA System on a Quantus Fluorometer (Promega, Madison, WI). The size and purity of the libraries were analyzed using the High Sensitivity D1000 Screen Tape on an Agilent 2200 TapeStation instrument. The libraries were pooled and run on an Illumina HiSeq 2500 sequencer using paired ends 100 bp Rapid Run format to generate 40 million total reads per sample.

After sequencing, raw reads were trimmed, removing low quality base, and were used in sequencing by Trimmomatic. The following criteria were applied for raw data cleansing: (1) Cut off when the average quality of sliding window (4-base wide) drops below 15; and (2) Reads shorter than 35 bp were discarded.

After the reads were aligned to the genome, Cuffquant was used on the resulting alignment files to compute the gene and transcript expression profiles. Cuffdiff, a part of the Cufflinks package, took the expression profiles and merged assemblies from two or more conditions to estimate the expression levels by calculating the number of RNA-Seq Fragments Per Kilobase of transcript per total Million (FPKM) fragments mapped. Cuffdiff tests the statistical significance of observed changes and identifies genes that are differentially regulated at the transcriptional or post-transcriptional level.

Cross-omics data were further analyzed by Perseus software by joining the data of proteome and transcriptome into one Perseus matrix. Both omics columns were sorted and transformed into ranks. A bivariant test was performed on each annotation term. Gene ontology (GO) enrichment analysis was performed to interpret sets of the genes with the functional characterization based on GO system of the classification. The potential interactions between the molecules were predicted by STRING.

### 11. Chromatin immunoprecipitation sequencing (ChIP-seq) and real-time quantitative chain reaction (CHIP-qPCR)

Mice ventricle tissue was cut into small pieces and homogenized in the homogenizer. Nuclei pallets were first extracted with the pre-extraction buffer of nuclear extraction kit (Abcam), then incubated in 0.8% formaldehyde at RT for 15 minutes to cross-link proteins to DNA. Glycine was added to a final concentration of 125 mM glycine to quench the formaldehyde and terminate cross-link reaction. Centrifugation was performed for 5 min, 4°C, 12000rpm, followed by the removal of cross-link buffer, the rinse of nuclei pellet with PBS, and centrifugation again as above. Supernatant was discarded. Pellet was resuspended, re in ChIP lysis buffer (50 mM HEPES-KOH pH7.5, 140 mM NaCl, 1 mM EDTA pH8, 1% Triton X-100, 0.1% Sodium Deoxycholate, 0.1% SDS, Protease Inhibitors), and incubated on ice for 15 minutes, then vortexed occasionally. Lysate was transferred to the special vial and sonicate (100% amplitude, on 30 seconds/ off 30 seconds) for 25 minutes to shear DNA to an average fragment size of 200 - 1000 bp. Pallet cell debris was centrifuged for 10 min, 4°C, 8000g. 50μl of this chromatin preparation was removed to serve as an input sample, and the remainder will be used for immunoprecipitation. Approximately 10μg of DNA per IP was recommended and each sample was diluted to 500μl with RIPA Buffer. Primary antibodies HDAC1 (1:250, GeneTex), Lamin B1 (1:200, GeneTex), H3K27ac (1:250, GeneTex) and H3K27me3 (1:250) were added to the indicated samples except the beads-only control and the sample was rotated at 4°C for 1 hour. The details of antibodies were listed in supplementary Table 1. Meanwhile, 25μl protein A/G beads (Thermo scientific) were used per IP, beads were washed three times in RIPA buffer. RIPA Buffer was aspirated, BSA was added to a final concentration of 0.1 μg/μl beads, and RIPA Buffer was added to double the bead volume. The sample was incubated for 30 minutes on rotator at RT. RIPA buffer was used to wash the sample once then it was added to double the bead volume. 50μl of blocked protein A/G bead was added to all samples and IP overnight with rotation at 4°C. The next day, the beads were collected with a magnetic stand and the unbound sample was removed. The following washes were performed, once in low salt wash buffer (0.1% SDS, 1% Triton X-100, 2 mM EDTA, 20 mM Tris-HCl pH 8.0, 150 mM NaCl), once in high salt wash buffer (0.1% SDS, 1% Triton X-100, 2 mM EDTA, 20 mM Tris-HCl pH 8.0, 500 mM NaCl), once in LiCl wash buffer (0.25 M LiCl, 1% NP-40, 1% Sodium Deoxycholate, 1 mM EDTA, 10 mM Tris-HCl pH 8.0). After each wash, the beads were collected with a magnetic stand and the supernatant was removed. DNA was eluted by adding 120μl of elution buffer (1% SDS, 100mM NaHCO3) to the protein A/G beads and vortexed slowly for 15 minutes at 30°C. The beads were collected with a magnetic stand and the supernatant was transferred into a fresh tube. 2μL RNase A (10 mg/mL) was added at 37°C for 30 minutes, then 2μL proteinase K (20 mg/mL) was added. The sample was then incubated at 55°C for 1 hour to reverse the crosslink. The eluted DNA fragment was purified with the Quick-DNA universal kit (Zymo Research) and diluted to 10ng/μL for DNA sequencing (by Illumina Miniseq) or the relative real-time quantitative chain reaction (by Illumina Eco) by the primers listed in Supplementary Table 2.

### 12. Absolute quantitative real-time PCR

The difference in the gene expressions of the cardiomyocyte treated with or without simvastatin was confirmed by absolute quantification method with Illumina Eco real-time PCR system. The primers were listed in Supplementary Table 3.

### 13. Immunoblotting

Mice ventricle tissue was cut into small pieces and homogenized in the homogenizer. Cytosol and nuclei lysates of mice ventricles or hiPSC-CMs were extracted with the nuclear extraction kit (Abcam, Cambridge, UK). Proteins were separated by 10% SDS-PAGE and transferred to a polyvinylidene difluoride (PVDF) membrane. The blot was blocked with 5% BSA and incubated with the indicated primary antibodies (Supplementary Table 1) at 4°C overnight. Then, the corresponding HRP-conjugated secondary antibodies were added (1:10000) at RT for 1 hour. The blot was incubated in SuperSignal West Femto Maximum Sensitivity Substrate (Thermo Scientific, CA, USA) for 5 minutes and exposed to UVP imaging system.

### 14. Chemicals

Dr. Chung-Ming Sun synthesized statin derivative KM-N1-029, which exhibited a purity exceeding 99% as confirmed by HPLC and NMR analysis. The salt form of the compound was dissolved in either distilled water or DPBS, while the non-salt forms were dissolved in DMSO. The other chemical reagents were purchased from Cayman Chemical (Michigan, USA).

### 15. Off-label use of simvastatin in the proband

Due to an awareness of the advantageous effects of simvastatin as revealed in the study conducted on cardiomyocytes derived from hiPSCs of the probands *in vitro* and on LVNC mice *in vivo*, the parents have requested an adjunctive therapy involving the off-label use of simvastatin for the proband. Subsequent to obtaining informed consent from her parents and verifying the normal levels of creatine kinase (CK), alanine aminotransferase (ALT), and aspartate aminotransferase (AST), the proband commenced a daily regimen of 5 mg (0.385 mg/kg/day) of simvastatin. Echocardiographic assessments and monitoring of plasma enzyme levels are to be conducted at each subsequent follow-up visit (ClinicalTrials.gov: NCT06632834).

### 16. Statistical method Data availability

A two-tailed paired or unpaired t test was employed to assess differences between two groups, while one-way ANOVA with Dunnett’s multiple comparisons test or Tukey’s post-hoc test was performed for comparisons involving samples from more than three groups.

## 17. Data availability

All data and materials used for supporting the findings of the current study are available from the corresponding authors upon reasonable request and with materials transfer agreements (MTAs) for animal models or human cells to any researcher in the aim of reproducing or extending the analysis.

## Results

### Patient characteristics and genetic information

The pedigree of this LVNC family is shown in Figure 1A. The infant (proband) (IIIb) initially presented with poor perfusion, hepatomegaly, cardiomegaly, and high level of NTproBNP (>3,5000 pg/mL). Echocardiogram performed under inotropic support showed dilated LV (the Z score of LV end-diastolic diameter, 4.7) with impaired LV contractility (LV ejection fraction, 26-30 % under milrinone 0.34 mcg/kg/min support) (Figure 1A). Family screening identified that her father (patient IIa) also had dilated LV and low LV ejection fraction (Figure 1A). Echocardiogram data of the other family members, including her grandfather (Ia), grandmother (Ib), mother (IIb), and sister (IIIa), were normal. Genetic examination of this LVNC family found that the proband and her father had heterozygous missense mutations in TNNT2 (chr1:201333464G>A; p.R141W) and MYPN (chr 10: 68210378 A>T; p.S1296T). Her father received carvedilol, losartan, and aspirin, and his cardiac status remained stationary with LVEF around 47-53%.

Besides from heart failure, there were no other extracardiac anomaly noted at presentation and during follow-up. CMR may offer more comprehensive information for myocardial pathology survey and is considered the gold standard for LVNC diagnosis. However, due to the young age and poor CMR resolution at that time, we opted for computed tomography instead of CMR for morphology and LVNC assessment. Computerized tomography in Figure 1B shows normal coronary arteries, dilated LV with hypertrabeculation and non-compaction of the proband (IIIb) heart (The compacted-to-compacted ratios at cardiac apex and lateral wall was 2.2 to 2.4). Myocarditis survey including viral isolation, cardiac enzyme, and ECG study was negative. Dilated cardiomyopathy with LVNC was diagnosed. During the subsequent follow-up, she received medications, including digoxin (3.5-4.5 mcg/kg/d), furosemide (2-3 mg/kg/day), captopril (0.8-1.2 mg/kg/day), carvedilol (0.15-0.3 mg/kg/day), and aspirin (2.5-4 mg/kg/day). About 2 years after the disease onset, her growth was slow, and the body weight was below the 3rd percentile. The LV ejection fraction by echocardiography remained poor, ranging from 17·5% to 24·5%. The NTproBNP was between 920 and 1200 pg/mL.

### Decrease of cell proliferation in human cardiomyocyte derived from the proband’s iPSC

EdU assay was conducted in human cardiomyocytes derived from iPSC of health mother (IIb) and the proband (IIIb) during day 12 to day14 after myogenic differentiation. Cardiomyocytes were identified by antibody against sarcomeric actinin α. Figure 1C shows the prominent decrease of myocyte proliferation in the early differentiated cardiomyocytes of LVNC group.

### TNNT2^R141W^ is the pathogenic mutation causeing cardiomyopathy

Mice harboring orthologous mutant genes, as identified in the probands, were employed to elucidate the deleterious impacts of the mutations. Distinctive features, such as a thinner non-compact myocardial layer and a thicker trabecular layer, were observed in H&E stained cardiac sections of E15.5 *Tnnt2*^R154W/+^ (T) mouse hearts, in contrast to those in wild type (WT) and *Mypn*^S1291T/+^ (M) mice (Figure 1D). A decrease in myocyte proliferation, as evidenced by a reduced number of myocytes expressing phospho-histone 3 (phospho-S10, PH3), was observed in embryonic E15.5 *Tnnt2*^R154W/+^ (T) mouse hearts in contrast to WT group (Figure 1E). Additionally, Figure 1F, depicting Masson’s Trichrome staining, reveals a dilated and thinner myocardium in mice carrying *Tnnt2*^R154W/+^ at 6 weeks of age without significant fibrosis. Assessing cardiac contractile function by echocardiography in mice carrying *Tnnt2*^R154W/R154W^ (TT) or *Mypn*^S1291T/S1291T^ (MM) at 6 weeks of age, Figure 1G demonstrates that mice carrying mutant *Tnnt*2 consistently exhibited marked heart failure.

### Loss of salt bridge coupling between TNNT2(R141W) and tropomyosin (E-257)

Figure 2A depicts the structure modeling of the protein interaction between troponin T (TnT) and tropomyosin (Tm) by Swiss Model. It highlights the substitution of Try(W) for troponin T Arg(R)-141 resulting in the disruption of the original salt bridge between troponin T (R-141) and tropomyosin (E-257). This structure alteration has impact on the movement of troponin T-tropomyosin during the transformation from calcium free to calcium bound state. As a result, it would potentially impair muscular contraction.

**Figure 2.**
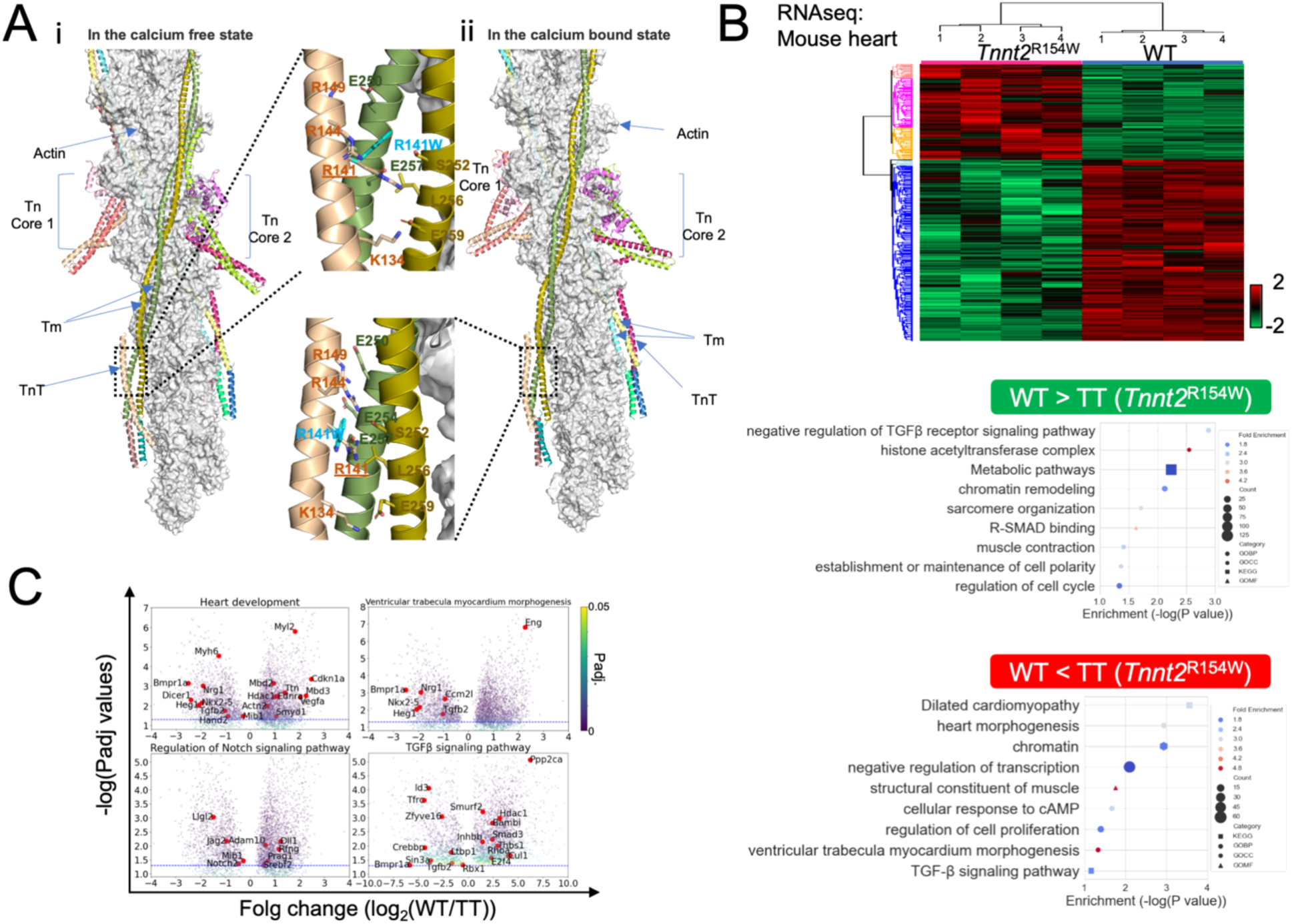
Mutant troponin T dose not only cause contractile dysfunction due to the loss of salt bridge between TNNT2(W141) and tropomyosin (E257) but also a significant transcriptomic alteration in *Tnnt*^2R154W/R154W^ mice hearts. (A) Close-up of residue R141 of TNNT2 (TnT) highlighting the rearrangement of the nearby electrostatic network in the absence (a) and presence (b) of Ca2+. The structures of human cardiac muscle thin filament in the absence and presence of Ca^2^+ have been determined by electron cryomicroscopy (PDB codes: 6KN7 and 6KN8). The guanidinium side-chain of TnT R-141 forms a salt bridge with the carboxylate side-chain of E-257 from tropomyosin (Tm) in the absence and presence of Ca^2+^. Substitution of Trp for Arg introduces a bulky aromatic side chain and disrupts polar contacts (hydrogen bonds or salt bridges). (B) RNA sequencing identified the statistically significant differential genes between the WT versus TT groups by unpaired t-tests. Hierarchical clustering heat maps were generated utilizing the Euclidean distance method to measure dissimilarities between sample pairs for each gene across all samples. The scale bar of Z-score was provided on the right side of the heatmap. Bubble plots were employed to visualize the biological functions or pathways associated with the differential genes of either higher level in WT or in TT, utilizing GOBP (circle), GOMF (triangle). GOCC (hexagon) and KEGG (square). The size of the symbols indicates the count number of the genes involved, while the color represents fold enrichment. (C) Volcano plots were utilized to depict the fold change of differential genes associated with specific KEGG pathways in comparison between WT and TT groups.

### Transcriptomic alteration in the hearts of Tnnt2^R154W/R154W^ (TT) mice

In addition to the disruption of a salt bridge between troponin T (W-141) and tropomyosin (E-257) in contribution to cardiac depression, Figure 2B shows that the altered transcriptomic profile of *Tnnt2*^R154W/R154W^ (TT) mice hearts could lead to the pathogenesis of cardiomyopathy. In TT mice hearts, genes related to the negative regulation of TGF-β signaling, sarcomere organization, metabolic pathways, cell polarity, and regulation of the cell cycle were downregulated. Conversely, genes associated with ventricular trabecula myocardium morphogenesis, the TGF-β signaling pathway, and the negative regulation of transcription were upregulated. Volcano plots in Figure 2C illustrate the significant changes in the expression of the genes involved in heart development, ventricular trabecula myocardium morphogenesis, regulation of Notch signaling pathways, and TGF-β signaling pathway. This highlights the additional pathogenic action of mutant TNNT2 in altering cardiac gene expressions.

### Decrease of nuclear TNNT2(R141W)-HDAC1 association

Liquid chromatography-tandem mass spectrometry (LC-MS/MS) was performed to identify the differential TNNT2-interaction-proteins pulldown by the fusion proteins of flag-SBP-TNNT2(WT) and flag-SBP-TNNT2(R141W) expressing in healthy- and LVNC-hCMs transfected with the plasmids of pDEST-*TNNT*2(WT) and pDEST-*TNNT*2(R141W) (maps in supplementary figure 3), respectively. It was found a decreased association between TNNT2(R141W) and HDAC1 and an increased association between TNNT2(R141W) and lamin B1 (LMNB1). In nuclear fraction of *Tnnt2*^R154W/R154W^ (TT) mouse hearts, Figure 3A shows that a significant reduction of mutant TNNT2 co-immunoprecipitated with HDAC1 in comparison to those of TNNT2(WT) group, yet the total protein expressions of HDAC1 and mutant TNNT2 were slightly decreased in *Tnnt2*^R154W/R154W^ (TT) mice hearts as shown in Supplementary Figure 4.

**Figure 3.**
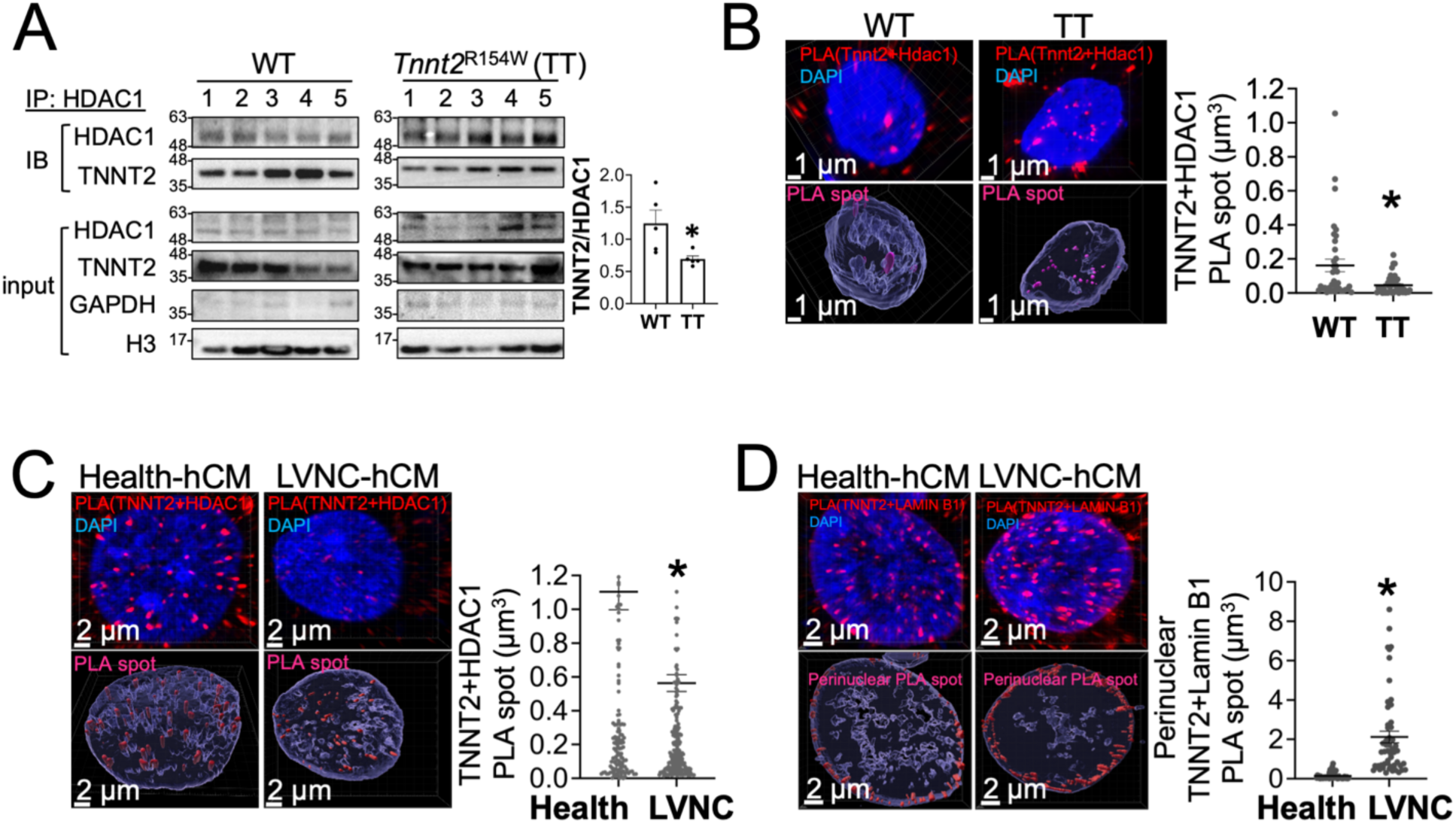
Mutant troponin T decreased the association with nuclear HDAC1 but increased the association with perinuclear LMNB1 (LAMIN B1) in LVNC cardiomyocytes. (A) The decrease of TNNT2 co-IP with HDAC1 from the nuclear fractions of *Tnnt2*^R154W/R154W^ (TT) mice hearts in comparison to that of wild type (WT) group. The level of co-IP TNNT2 was normalized to that of IP-HDAC1 in each sample. Data are mean±SEM (n=5). *P<0.05, versus WT group, by unpaired t test. Proximity ligation assay (PLA) was employed to identify the colocalized spots of TNNT2 & HDAC1 (red PLA spot: troponin T (TnT)+HDAC1; blue: DAPI) in the nuclei of (B) WT & TT mice ventricular cardiomyocytes and (C) Health-hCM & LVNC-hCM. The lower panels under cytofluorescence images show the contour of PLA spots (pink) in myocyte nuclei (purple) illustrated by Imaris surface software. The volume of PLA spot was quantified by Imaris surface. The scatter plot of PLA spot volume was illustrated in the right panel. Data are mean±SEM (n=3, three different samples in each group). *P<0.05, versus WT or health group, by unpaired t test. (D) Comparison of the volume and the perinuclear distribution of PLA spot constructed by antibodies targeting TNNT2 and LMNB1 (red PLA spot: troponin T (TnT)+ lamin B1; blue: DAPI) in Health- and LVNC-hCMs via the image analysis by Imaris surface. TNNT2-lamin B1 (LMNB1) complex distributed within the border of nuclei DAPI was illustrated as the pink perinuclear PLA spots. The scatter plot of the perinuclear PLA spot volume was illustrated in the right panel. Data are mean±SEM (n=3, three different batches of hCMs in each group). *P<0.05, versus health group, by unpaired t test.

A proximity ligation assay (PLA) was performed to corroborate the reduced interaction between mutant TNNT2 and HDAC1 in the intranuclear locale of TT mouse cardiomyocytes (Figure 3B), as well as the attenuated association between mutant TNNT2 and HDAC1 in human cardiomyocytes derived from the proband’s hiPSC (LVNC-hCM) (Figure 3C), respectively. PLA spot was visualized and spot volume was quantified using Imaris Surfaces, as shown in the right panels of Figure 3B-C. Furthermore, an increased association between mutant TNNT2 and lamin B1 (LMNB1) was observed in LVNC-hCM (Figure 3D), especially the increased volume and number of PLA spots in the perinuclear regions of LVNC-hCM quantified by Imaris Surfaces.

### Attenuated intranuclear TNNT2(R141W)-HDAC1 association caused cardiac epigenetic perturbation and the dysregulation of cardiac gene expressions

To further elucidate the differences in chromatin H3K27ac profiling between wild-type (WT) and *Tnnt2*^R154W/R154W^ (TT) mouse hearts, chromatin immunoprecipitation followed by high-throughput sequencing (ChIP-seq) was performed. Figure 4A shows the percentage of the significantly altered H3K27ac profiling in the exons, the gene promoter regions, introns, and the others. There are 458 common genes with significant changes in both H3K27ac profiling in the gene promoter regions by ChIPseq and the transcription levels by RNAseq (Figure 4B). The heat map presented in Figure 4C illustrates the hierarchical heat map of the genes with significant change in H3K27ac marks within the promoter regions. The top MSigDB gene-sets that significantly enriched (FDR < 0.05) in genes exhibiting decreasing or increasing H3K27ac modifications in *Tnnt2*^R154W/R154W^ (TT) mice hearts are depicted in the upper & lower panel of Figure 4D, respectively. Figure 4E shows a notable decrease in H3K27ac marks within cardiac muscle genes (*Actc1*, *Actn2*, *Lmod2*, *Tnnt2*), Notch signaling genes (*Dll4*, *Hey2*), and chromatin modifier (*Hdac1*) in *Tnnt2*^R154W/R154W^ (TT) mouse hearts. Alternatively, a marked increase was also found in H3K27ac marks within TGFβ signaling genes (*TGFβ*1, *TGFβ*2, *TGFβr*1, *Id3*), Notch signaling genes (*Notch4*, *Hes1*), and chromatin modifier (*Ezh*2) as shown in Figure 4F. It was further found a notable reduction of HDAC1- and LMNB1 (lamin B1)-marks within the promoter region of *Tgfβ*1 (Supplementary Figure 5) and *Ezh*2 (Supplementary Figure 6A). Furthermore, the upregulation of EZH2 in *Tnnt*2^R154W/R154W^ (TT) mice hearts (Supplementary Figure 6B) consequently led to the increase of H3K27me3 marks in *Dll4* promoter region in *Tnnt*2^R154W/R154W^ (TT) mice hearts (Supplementary Figure 6C). Similar finding of the upregulation of EZH2 was validated in LVNC-hCMs (Supplementary Figure 6D). Further study demonstrated that GSK503 (0.3 μM, a selective EZH2 inhibitor) can significantly increase *DLL*4 expression in LVNC-hCM in Supplementary Figure 6E.

**Figure 4.**
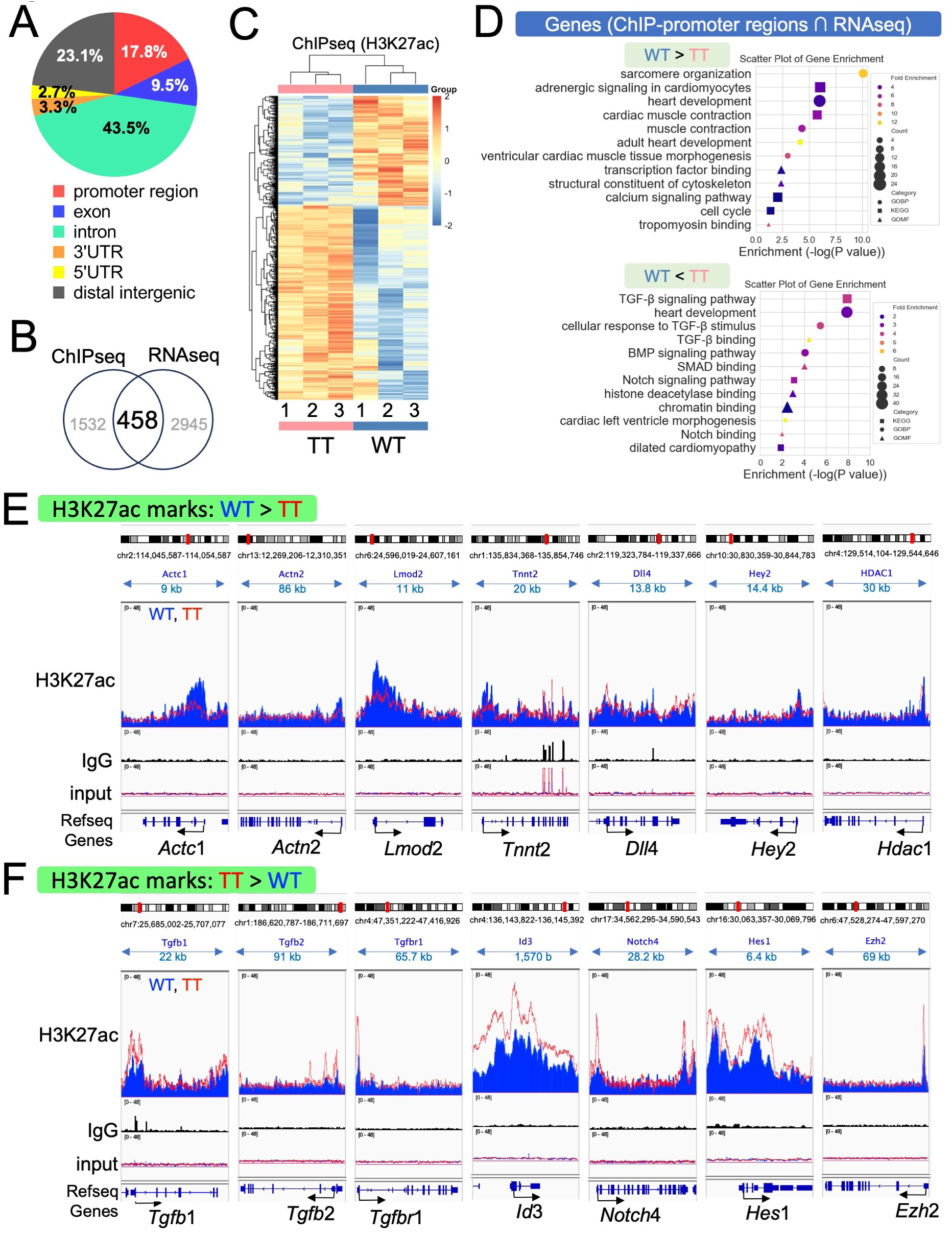
Mutant troponin T altered H3K27ac marks in cardiac genes involving in heart development and cardiac muscle contraction. (A) The percentage of H3K27ac marks in the different chromosome regions examined by ChIPseq. (B) There are 458 genes that exhibit consistent change in both H3K27ac marks in the gene regions and the transcriptional expressions. (C) Hierarchical clustering heatmap of mice genes exhibiting significant change in gene regions where H3K27Ac marks were identified by ChIP-seq in the hearts of wild type (WT) and *Tnnt2^R^*^154^*^W^* (TT) mice (three different mice hearts in each group). (D) The bubble plots reveal the top enriched MSigDB gene-sets of 458 genes with significant decrease (upper panel) and unique increase (lower panel) of H3K27ac modification in gene regions of TT group. (E-F) ChIP-seq tracks for H3K27ac at the regions of representative genes with loss of H3K27ac marks ( *Actc2*, *Actn2*, *Lmod2*, *Tnnt2*, *Dll4*, *Hey2*, *Hdac1*) and gain of H3K27ac (*Tgf*β1, *Tgf*β2, *Tgf*β*r*1, *Id3*, *Notch*4, *Hes*1, *Ezh*2) in TT group (red track) in comparison to those in WT group (blue track). All data were normalized to the corresponding input of each sample. The representative trace was summed from three independent samples in each group. Chromosomal loci & reference sequence of these genes was shown in the top & bottom panels, respectively.

### Excessive TGF-β stimulation and reduced Notch activity inhibited LVNC-hCM proliferation

EdU assay was performed to measure the change of LVNC-hCM proliferation *in vitro*. Figure 5A shows the inhibitory effect of TGF-β1 on cell proliferation of healthy human cardiomyocytes (health-hCM). However, the absence of a discernible inhibitory response to TGF-β1 on cell proliferation in LVNC-hCM, coupled with a pronounced increase in myocyte proliferation in the presence of A83-01, a TGF-βR1 inhibitor, indicated excessive TGF-β stimulation under basal conditions in LVNC-hCM.

**Figure 5.**
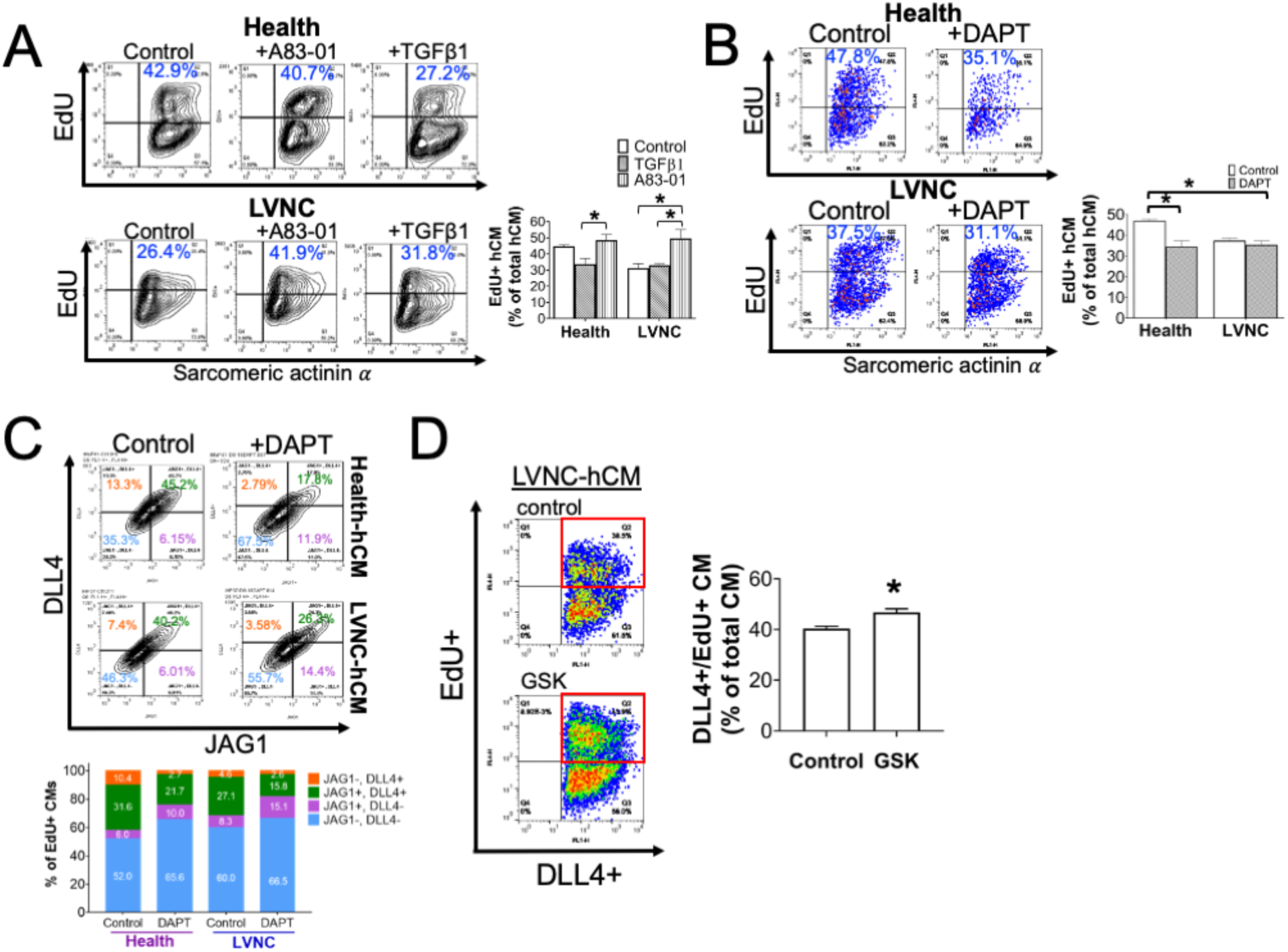
The excessive TGFβ stimulation and the reduced NOTCH activity in LVNC-hCM caused cardiac growth defect. Pulse-chase EdU assay with Alexa Fluor 647 fluorescent label was performed to examine cell proliferation of hiPSC-derived cells with the indicated drug treatment during day 12 through day 16 after myogenic differentiation. Human cardiomyocytes were identified by mouse monoclonal antibody targeting sarcomeric actinin α (SA, with anti-mouse IgG secondary antibody conjugated with Alexa Fluor 488). Flowcytometry was performed to quantify the cell population by BD LSRFortessa. (A) The proliferation rate of LVNC-hCM was lower than that of Health-hCM. TGFβ1 (3 nM) can prominently decrease EdU+ myocytes in health-hCM but not those in LVNC-hCM. A83-01 (3 μM), a TGFβR1 inhibitor, can markedly increase EdU+ myocytes in LVNC-hCM. Data are mean±SEM (n=3 in each group). *P<0.05, versus control or the indicated groups by one-way ANOVA with post-hoc Tukey’s test. (B) The difference of EdU+/SA+ myocyte population between control and DAPT (0.3 μM)-treated groups was defined as the capacity of Notch-dependent myocyte proliferation that was significantly decreased in LVNC-hCM. Data are mean±SEM (n=3 in each group). *P<0.05, versus control by one-way ANOVA with post-hoc Tukey’s test. (C) DLL4+ myocytes were decreased in LVNC-hCM and could be further depleted by DAPT (0.3 μM). The lower panel shows the cell population percentage averaged from three independent samples (n=3 in each group). (D) The cell population of DLL4+/EdU+ LVNC-hCM (in red square) can be increased by GSK (0.3 μM). Data are mean±SEM (n=4 in each group). *P<0.05, versus control by unpaired t test.

In addition, Figure 5B shows that myocyte proliferation can be significantly attenuated by DAPT, a γ-secretase inhibitor, in healthy-hCM but not in LVNC-hCM, implying a reduction of Notch-dependent cell proliferation in LVNC-hCM. This observation was further substantiated by the notable decrease in DLL4+ myocytes, rather than JAG1+ myocytes in LVNC-hCM as depicted in Figure 5C. The functional validation in Figure 5D underscores the substantial repopulation of DLL4+/EdU+ LVNC-hCM achieved through treating GSK503 (a EZH2 inhibitor) that can recover DLL4 expression (Supplementary Figure 6E).

### Simvastatin can rescue TNNT2(R141W)-HDAC1 association to recover cardiac epigenetic, proteomic profile and function

Statins have previously been documented to possess anticancer properties by modulating cancer epigenetics through their interaction with HDAC^19, 20^. A repurposed drug screen was performed to identify simvastatin being capable of acting as a chemical chaperone, effectively restoring the association between TNNT2(R141W) and HDAC1 in a dose-dependent manner within LVNC-hCM transfected with pDEST-flag-SBP-TNNT2(R141W), as depicted in Figure 6A. The reduction of TNNT2 co-immunoprecipitated with HDAC1 from the nuclear fraction of LVNC-hCM can be substantially reestablished in the presence of simvastatin at 0.1 μM, as demonstrated in Figure 6B. This is in line with the concurrent attenuation of EZH2 expression in simvastatin-treated LVNC-hCM, as shown in Figure 6C. Furthermore, a dose-dependent effect of simvastatin in the increase of LVNC-hCM proliferation was observed, as presented in Figure 6D. Simvastatin treatment from day 30 through day 70 can also dose-dependently preserve the intracellular calcium handling properties measured by continuously optical pacing at 1Hz in LVNC-hCM expressing GECIs and ChR2-K-GECO, as indicated in Figure 6E.

**Figure 6.**
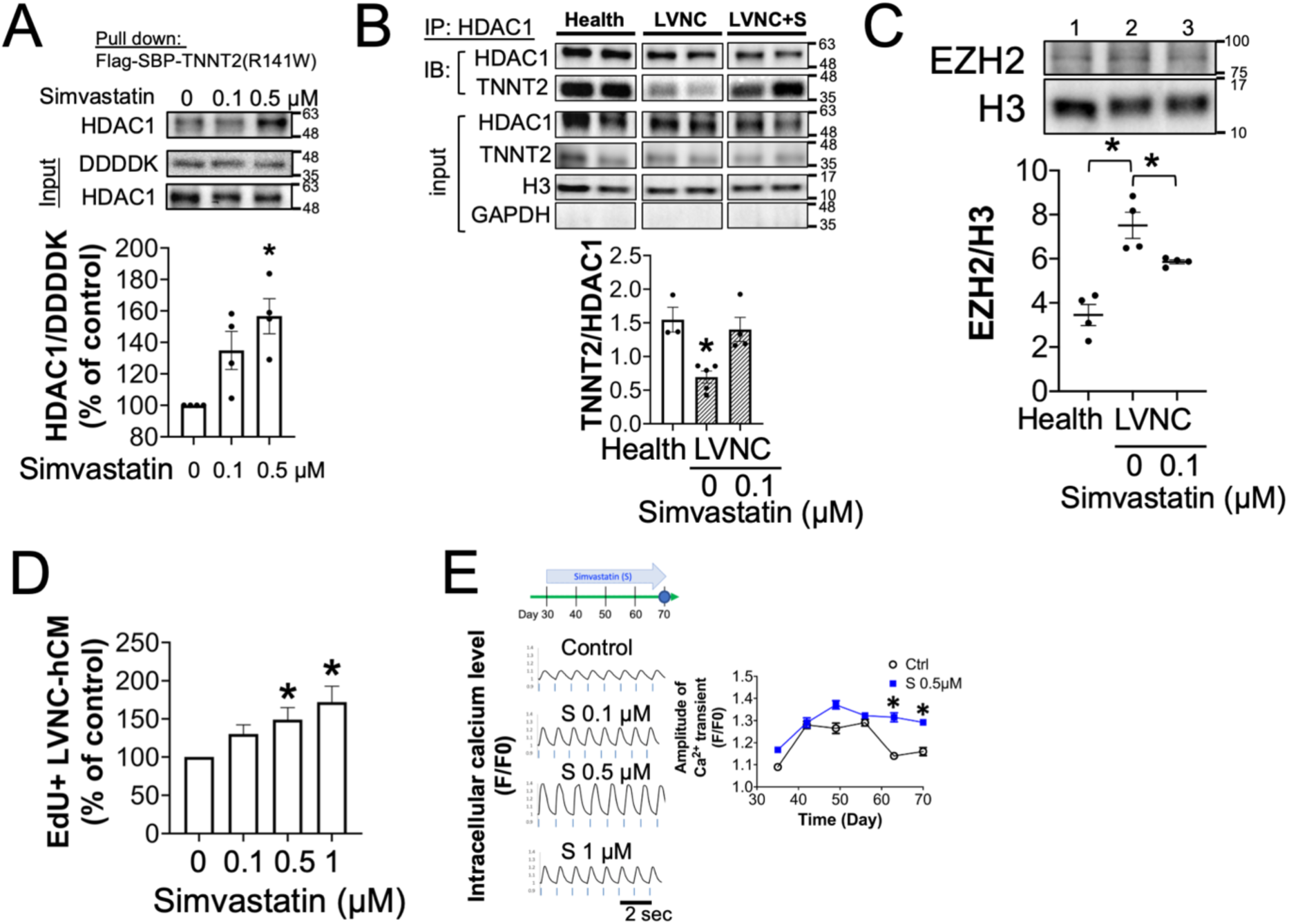
Simvastatin can restore the association between TNNT2(R141W) and HDAC1 to improve cardiac growth and function of LVNC-hCM. (A) TNNT2(R141W)-associated proteins were pulled down by streptavidin-beads from nuclear fraction of LVNC-hCM transfected with pDEST-TNNT2(R141W) encoding the fusion protein of flag-SBP-TNNT2(R141W) in the absence or presence of simvastatin at the concentrations of 0.1 and 0.5 μM. Simvastatin can dose-dependently increase the associated TNNT2(R141W)-HDAC1 complex. Data are mean±SEM (n=4 in each group). *P<0.05, versus control by one-way ANOVA with post-hoc Tukey’s test. (B) Co-immunoprecipitation of HDAC1 and TNNT2 from nuclear fraction of LVNC-hCM treated with or without simvastatin (0.1 μM). A significant increase of TNNT2 co-immunoprecipitated by HDAC1 was observed in simvastatin-treated LVNC-hCM. Data are mean±SEM (n=3-5 in each group). *P<0.05, versus LVNC by one-way ANOVA with post-hoc test by Dunnett’s multiple comparisons test. (C) The upregulation of EZH2 in LVNC-hCM was prominently abolished by simvastatin (0.1 μM). Data are mean±SEM (n=4 in each group). *P<0.05, versus LVNC without simvastatin treatment by one-way ANOVA with post-hoc Tukey’s test. (D) Click-it EdU assay with Alexa Fluor 647 fluorescent label was performed to examine the proliferation of myocytes identified by mouse monoclonal antibody targeting sarcomeric actinin α with secondary antibody conjugated with Alexa Fluor 488. Simvastatin can dose-dependently increase pulse-chase-labeled EdU+ LVNC-hCM during day 12-16 after myogenic differentiation at the concentrations ranging from 0.1, 0.5 and 1 μM. Data are mean±SEM (n=3 in each group). *P<0.05, versus control by one-way ANOVA with post-hoc Tukey’s test. (E) LVNC-hCMs expressing K-GECO (a genetically encoded calcium indicator) and channelrhodopsin-2 delivered by AAV were used to measure intracellular calcium transients elicited by optical pacing at 1 Hz (indicated by blue lines). Intracellular calcium transients were measured every week. A marked decrease in the amplitude of calcium transients of LVNC-hCMs was observed from day 50 through 70. The representative traces in left panel show a significant increase of calcium transients in simvastatin (S)-treated group at the concentrations of 0.1, 0.5, and 1 μM in LVNC-hCM from day 30 through day 70 after myogenic differentiation. Data are mean±SEM (n=10 different LVNC-hCMs in control, n=22 different LVNC-hCMs in S group). *P<0.05, versus control at the same time point by unpaired t test.

βAR-stimulated positive inotropic effect was prominently blunted in LVNC-hCM *in vitro* (Figure 7A). Subsequently, an investigation was carried out to assess whether HDAC inhibitor or other statins could exert a therapeutic effect in recovering the responsiveness to βAR-stimulation in LVNC-hCM *in vitro*. The pan HDAC inhibitor SAHA demonstrated a mild improvement in the β-responsiveness of LVNC-hCM as displayed in Figure 7A. Furthermore, only simvastatin and lovastatin, characterized by the inclusion of the essential structure comprising naphthalene and mevalonolactone (Supplementary Figure 7), exhibited a significant beneficial effect in enhancing β-responsiveness in LVNC-hCMs following treatment from day 30 to 70, as shown in Figure 7A. Statins lacking naphthalene, such as atorvastatin and fluvastatin, or statin derivatives lacking the hydroxy side group on mevalonolactone, such as KM-N1-029, did not manifest any beneficial effects in muscular contraction and β-responsiveness in LVNC-hCMs, as illustrated in Figure 7A. Proteomic study further demonstrated that simvastatin can restore the protein profiles of LVNC-hCMs (iH & iF) approaching to those of health-hCMs (iM & health.m) in terms of promoter activity, cardiac muscle, and metabolism constructed the network by STRING with GOBP (Gene Ontology Biological Process) annotations, as illustrated in Figure 7B.

**Figure 7.**
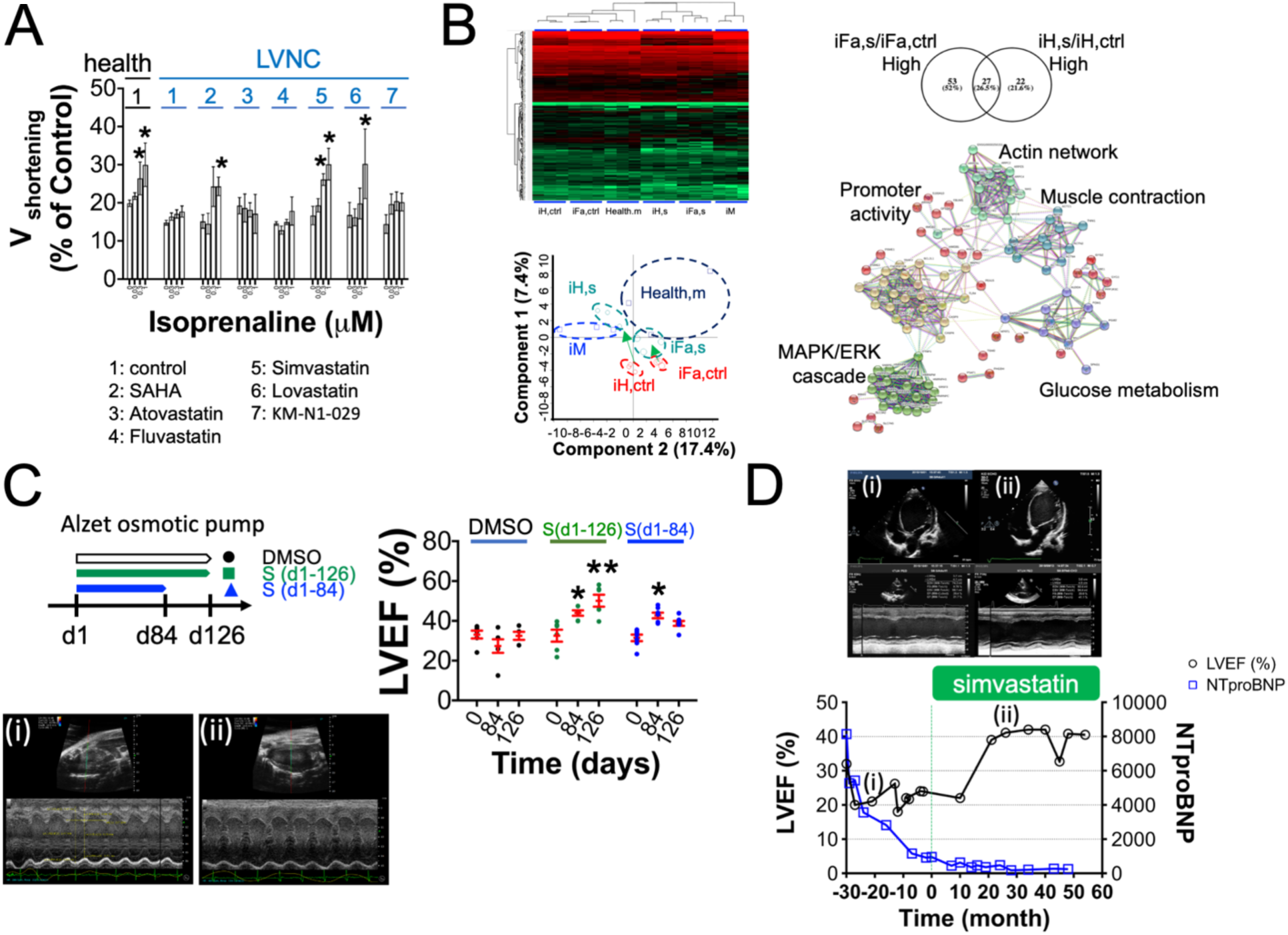
Functional improvement of LVNC cardiomyocytes *in vitro* and hearts *in vivo* by simvastatin. (A) The structure and activity relationship between simvastatin & its analogs was established through measuring myocyte shortening velocity (V_shortening_) in response to isoprenaline (at the concentrations of 0.03, 0.3 and 1 μM) in LVNC-hCMs treated with the indicated reagents (1: control, 2: SAHA (200 nM), 3: Atorvastatin (300 nM), 4: Fluvastatin (300 nM), 5: Simvastatin (300 nM), 6: Lovastatin (300 nM) and 7: KM-N1-029 (300 nM). Health-hCM was used as normal control in response to β-stimulation. (B) Heatmap and PCA of proteomic profile of health-hCM (iM & Health,m)- and LVNC-hCM (iFa & iH) on day 70 after the treatment with vehicle (ctrl: control) or with simvastatin (S, 0.1 μM) from day 30 through day 70. The functional protein association network of simvastatin-recovered proteins expressing in both of iFa & iH LVNC-hCMs within the clusters of GOBP annotations including muscle contraction, glucose metabolism and promoter activity was constructed using STRING. The green arrows indicate the shift direction of the proteomic profile in two LVNC-hCM lines with simvastatin-treatment. (C) LVEF of Tnnt2^R154W/+^::Mypn^S1291T/+^ mice could be prominently improved in continuous simvastatin treatment via osmotic pump (165 nmole/mouse/day) for consecutive 126 days. Severe heart failure was relapsed on day 126 once withdrawal simvastatin from day 84. Data are mean±SEM (n=6 in each group). *P<0.05, versus LVEF before treatment (day 0) by paired t test. (D) The change of LVEF (circle) and plasma NTproBNP (square) of this proband before and after simvastatin (5 mg per day, p.o.) treatment. The upper panel is the representative pictures of echocardiography at the time points of (i) two years before simvastatin treatment and (ii) two years after consecutive treatment with simvastatin.

### Therapeutic effect of simvastatin in LVNC mice in vivo

*Tnnt2*^R154W/+^::Mypn^S1291T/+^ mice were utilized to assess the therapeutic potential of simvastatin *in vivo*. A prolonged therapeutic regimen was implemented, involving the use of an osmotic pump (ALZET, model 2006, total duration: 6 weeks) for controlled drug delivery to *Tnnt2*^R154W/+^::Mypn^S1291T/+^ mice *in vivo*. The implanted osmotic pump was replaced with a new one every 6 weeks. Continuous treatment with simvastatin (165 nmole/mouse/day) led to a gradual and substantial improvement in left ventricular ejection fraction (LVEF) in *Tnnt2*^R154W/+^::Mypn^S1291T/+^ mice, as observed over a 126-day period (Figure 7C). However, upon discontinuation of the drug after day 84, a relapse in the progression of heart failure was noted.

### The therapeutic benefit of off-label use of simvastatin in TNNT2^R141W/+^-LVNC patient

Due to the evidence of simvastatin-produced beneficial effects in LVNC-hCM *in vitro* and LVNC-mice *in vivo*, the off-label use of simvastatin in add-on manner with the state-of-the-art medications was initiated after having the patient’s informed consent. The proband received 5 mg once daily (0.385 mg/Kg/day). No adverse effects were observed. The echocardiography of her cardiac condition in Figure 7D shows (i) LVEF of 21.7% on the time point of two years before simvastatin treatment and (ii) the improved cardiac function to LVEF of 41.1% after the simvastatin therapy for consecutive two years (ClinicalTrials.gov: NCT06632834). Her body weight improved from the 3^rd^ percentile to the 10^th^ percentile. The NTproBNP level was lowered to 182.6 pg/mL in concurrence with the decrease of cardiac size (Z score 3.93) measured by echocardiography. No adverse effects were observed during treatment.

## Discussion

Given the documented existence of over 32 distinct genetic mutations associated with Left Ventricular Non-Compaction Cardiomyopathy (LVNC) ^21^, there arises an opportunity for pharmaceutical interventions to selectively target the pathological signaling cascades induced by these mutant genes. From investigating the pathological mechanism of a missense mutation in TNNT2, the present study provided novel insights into the epigenetic regulatory function of intranuclear troponin T (TNNT2) through serving as a sponge of HDAC1 for the maintenance of cardiac epigenetic to facilitating the preservation of cardiac functional gene expressions. In addition, TNNT2(R141W) exhibited a diminished affinity to intranuclear HDAC1 and an enhanced affinity to lamin B1, especially perinuclear distribution, in LVNC myocytes. The redistributed intranuclear HDAC1 resulted in a decrease in H3K27ac marks in specific cardiac genes associated with muscle contraction, calcium handling, and mitochondrial metabolism, consequently leading to gene downregulation and cardiac dysfunction. Alternatively, the decrease of HDAC1 associated with TNNT2(R141W)-lamin B1 complex in perinuclear region may contribute to an increase in H3K27ac marks in the genes involved in TGFβ signaling and EZH2. It consequently leads to the inhibition of cardiac growth via TGFβ-overstimulation and the attenuated NOTCH activity due to DLL4 downregulation caused by the overexpressed EZH2-mediated increase of H3K27me3 mark in DLL4 promoter region. Through drug repurposing screen in hiPSC-derived cardiomyocytes from affected individuals (LVNC-hCMs) to explore the chemical chaperone for reinstating TNNT2(R141W)-HDAC1 association, simvastatin was identified to exert therapeutic potential capable of improving cardiac function by normalizing cardiac epigenetics and omics profiles. The therapeutic efficacy of simvastatin was demonstrated in both LVNC-hCMs *in vitro* and LVNC mice carrying Tnnt2^R154W^ *in vivo*. Continuous simvastatin treatment was found to be necessary to prevent the recurrence of heart failure progression, as evidenced by the *in vivo* LVNC mouse study. This study successfully elucidated the role of intranuclear TNNT2 in cardiac epigenetic homeostasis, the underlying mechanism of TNNT2(R141W) in LVNC pathogenesis, and the potential druggable targets in the signaling cascades.

Different mutation sites on *TNNT*2 have been associated with various types of cardiomyopathy (CMP). Mutations in *TNNT*2 on the sites of I79N^22^, R92L/R92Q^23^/R92W^24, 25^, F110I, R130C^24^, Delta E160^26^, E163K^22^, K263R^27^, and R278C^28^ have been reported to be linked to familial hypertrophy cardiomyopathy (HCM). In contrast, mutations including N83H^29^, L84F^30^, R141W^22^, R173W^22, 31^, R183W^32^, and DeltaK210^33^ have been associated with dilated cardiomyopathy (DCM). However, the precise mechanisms by which different *TNNT*2 mutations lead to HCM or DCM, as well as the development of therapeutic medications targeting the pathological signaling pathways caused by these mutations, are not yet fully understood. In the present study, *TNNT2*^R141W^ mutation was identified from an LVNC infant with DCM feature. Prominent hypertrabeculation and left ventricle noncompaction was also observed in the embryonic hearts of the knock-in mice harboring the orthologous *Tnnt2*^R154W^ mutation. *TNNT*2^R141W^ has been reported to be associated with DCM.^22^ In the present study, DCM features appeared in adult *Tnnt2*^R154W^ mice at the age over 6 weeks (Figure 1F), suggests that LVNC with *TNNT2*^R141W^ mutation can further progress to DCM. This suggests that the phenotypic manifestation of LVNC is intricately determined by distinct genetic mutations within each individual and the timing of disease surveillance. This underscores the vital significance of employing advanced cardiac imaging techniques in pediatric patients to ensure precise diagnosis, thus establishing a solid foundation for the progression of precision medicine in the future.

Computer simulations of the protein structure have predicted that the mutant troponin T (TNNT2(R141W)) may lose its functional coupling with tropomyosin due to the disruption of the salt bridge between TNNT2(W-141) and the carboxylate side-chain of tropomyosin (E-257). This molecular alteration could potentially result in muscle weakness, which cannot be rescued by simvastatin. However, manipulating TNNT2(R141W)-mediated pathological signaling should be promising to control disease progression. Prior studies have suggested the nuclear localization of TNNT2 in cardiomyocytes.^31, 34^ In TNNT2(R173W)-induced dilated cardiomyopathy (DCM), it is proposed that an increased interaction of nuclear TNNT2(R173W) with KDM1A (LSD1) and KDM5A (JARID1) might induce cardiac epigenetic modifications by redistributing these histone demethylases, thereby increasing H3K4me3 marks in the regions of PDE2A. These modifications in turn could lead to an abnormal upregulation of phosphodiesterase (PDE2A) and contribute to the loss of β-AR responsiveness in DCM.^31^ The current study further investigated the involvement of nuclear TNNT2 in the formation of the TNNT2-HDAC1 complex (Figure 3). This complex is suggested to serve as an HDAC1 sponge, regulating the intranuclear distribution of HDAC1 and maintaining the euchromatin structure of functional cardiac genes, thereby facilitating cardiac gene expressions. Notably, the mutant TNNT2(R141W) exhibited a diminished interaction with intranuclear HDAC1 while displaying an increased association with lamin B1 in the perinuclear region. Consequently, the redistribution of HDAC1 from intranuclear TNNT2(R141W) may contribute to the formation of a heterochromatin structure in cardiac genes responsible for encoding functional proteins involved in muscle contraction and heart development (Figure 4D-F). Conversely, an enhanced euchromatin state may be observed in genes related to TGFβ signaling molecules and EZH2. Notably, early differentiated LVNC-hCMs displayed excessive TGFβ activity, which inhibited myocyte proliferation (Figure 5A), leading to cardiac growth impairment (Figure 1). Additionally, the aberrant upregulation of EZH2 induced by TNNT2(R141W) also contributed to a defect in cardiac growth. This defect arises from the reduction of Notch-dependent stimulation in myocyte proliferation, occurring through the inhibition of DLL4 expression via epigenetic modifications. Previous reports have documented the overexpression of EZH2 in ischemic cardiomyopathy^35^ and dilated cardiomyopathy.^36^ EZH2, as an enzymatic component of the polycomb repressive complex 2 (PRC2) belonging to the SET domain family of histone-lysine N-methyltransferases, facilitates the trimethylation of lysine-27 on histone H3 (H3K27me3), thereby promoting heterochromatin formation and transcriptional repression.^37^ The observed increase in H3K27me3 marks and decrease in H3K27ac marks in the *DLL*4 gene correspond to the aberrant upregulation of *EZH*2 and the downregulation of *DLL*4 in LVNC-hCM. Functional validation demonstrated that the direct inhibition of EZH2 significantly enhances DLL4 expression in LVNC-hCM, ultimately leading to improved myocyte proliferation (Figure 5D).

The schematic diagram I illustrates the druggable targets that have been identified in the pathological signaling cascade mediated by TNNT2(R141W). Given the reduced interaction of nuclear TNNT2(R141W) with HDAC1, resulting in an epigenetic perturbation caused by the redistributed HDAC1, one noteworthy target emerges as HDAC1. It has been reported that HDAC inhibition may improve cardiac diastolic dysfunction^38, 39^. Notably, our study found SAHA, a pan HDAC inhibitor, exhibits promise in partially restoring β-AR responsiveness by rectifying heterochromatin patterns in cardiac functional genes that have been impacted by the redistributed HDAC. However, the increased TNNT2(R141W)-laminB1 association concomitantly with the diminished TNNT2(R141W)-HDAC1 association within the perinuclear region leads to anomalous euchromatin configurations in genes such as TGFβ-signaling molecules and EZH2. HDAC inhibitors might worsen this pathological signaling. Thus, the utilization of a TGFβR1 inhibitor (A83-01) appears to counteract TGFβ-mediated inhibitory influences on cell proliferation, thereby enhancing LVNC myocyte proliferation, as visually depicted in Figure 5A. Within the context of restoring DLL4 expression, employing an EZH2 inhibitor (GSK503) has demonstrated a significant propensity to increase LVNC-hCM proliferation. Although these approaches can improve cardiac growth, they cannot prevent heart failure progression caused by TNNT2(R141W)-induced HDAC redistribution. An ideal pharmacological strategy for comprehensively obstructing TNNT2(R141W)-mediated pathological pathways entails the restoration of interaction between TNNT2(R141W) and HDAC1. Simvastatin, a hit repurposed drug derived synthetically from a fermentation product of Aspergillus terreus, can reestablish the TNNT2(R141W)-HDAC1 association and significantly improve cardiac function among statins. The potential direct interaction between statins and HDAC2 was reported by computation modeling^20^. The present study further demonstrated that only simvastatin and lovastatin, rather than the other statins shown in supplementary Figure 7, can act as a chemical chaperone to improve cardiac growth and function of LVNC-hCMs *in vitro* and *Tnnt2*^R154W/+^::Mypn^S1291T/+^ mice *in vivo* through reinstating the association of TNNT2(R141W) and HDAC1. Although the dosage range and specific indications remain unclear, our study shows the optimal dose of simvastatin, 165 nmol/mouse/day, via subcutaneous-osmotic-pump-administration can significantly produce functional improvement in *Tnnt2*^R154W^ mice *in vivo* (Figure 7C).

Simvastatin has been used in children with various underlying diseases, including a history of organ transplantation, renal disease, and familial hypercholesterolemia^40, 41, 42^. No significant safety concerns have been identified. Because of the beneficial effects of simvastatin from the study in cardiomyocytes derived from the probands’ hiPSCs *in vitro* and in LVNC mice *in vivo*, an add-on therapy with off-label use of simvastatin in the proband was initiated after obtaining her parents’ agreement. The baseline of the proband’s cardiac condition during 30 months after the disease onset shows the poor prognosis of the proband’s cardiac function (LVEF 20-30 %) measured by echocardiography and NTproBNP level above 900 pg/mL. The proband began to receive 5 mg (0·385 mg/Kg/day) once daily (ClinicalTrials.gov: NCT06632834). After the simvastatin therapy for consecutive 2 years, cardiac function was significantly improved to be LVEF above 41.1%. Her body weight improved from the 3^rd^ percentile to the 10^th^ percentile. The NTproBNP level was lowered to 182.6 pg/mL in association with the decrease of cardiac size (Z score 3.93). No adverse effects were observed during the treatment.

These observations highlight the potential for epigenetic modulation as a therapeutic strategy for LVNC and underscore the importance of further investigations to elucidate the underlying mechanisms driven by different TNNT2-mutations mediated epigenetic dysregulation. Ultimately, a better understanding of these processes may contribute to the development of innovative therapeutic approaches for patients with LVNC. This preclinical trial in a dish *in vitro* and functional validation in LVNC mice *in vivo* provide a warrant of success for the future clinical trial.

## Acknowledgments

We thank the grant support provided by National Science and Technology Council in Taiwan (NSTC), National Health Research Institutes (NHRI), and the Excellent Translational Medicine Research Projects of National Taiwan University College of Medicine and National Taiwan University Hospital (ETMRP).

NSTC105-2320-B-002 -036 -MY3, NSTC106-2314-B-002-207, NSTC108-2320-B-002 -027 - MY3, NHRI-EX112-11141SI, ETMRP-106R39012 & -1105F025-02 to C.W.P.

NSTC106-2314-B-002-207 & NSTC109-2314-B-002-140 to W.M.H.

NSTC-105-2314-B-002 -109 -MY3, NSTC107-2319-B-001 -003, ETMRP-106R39012 and ETMRP-1105F025-02 to H.H.N.

We would like to acknowledge the service provided by Hua-Man Hsu & Shao-Chun Hsu at the imaging core facility of the First Core Laboratory, and Hsueh-Chin Chen & Wei-Jou Lin at Laboratory Animal Center, National Taiwan University College of Medicine. We thank the technical services provided by Suz-Ying Chen in the maintenance of hiPSC, the technical service for the generation of gene-targeting knock-in mice by the Transgenic Mouse Model Core Facility of the National Core Facility Program for Biotechnology, Ministry of Science and Technology, Taiwan and the Gene Knockout Mouse Core Laboratory of National Taiwan University Center of Genomic Medicine, Transmission Electron Microscopy (TEM) service by Dr. Hsiuni Kung at Graduate Institute of Anatomy and Cell Biology & Mr. Yansheng Wu at College of Medicine, Fu Jen Catholic University, and ChIPseq by Chia-Lang Hsu at Department of Medical Research in NTUH. We thank the staff of the Sequencing Core and the High-throughput Genomics and Big Data Analysis Core, Department of Medical Research, National Taiwan University Hospital for technical support. We thank the English editing by Sen-Han Chang

## Author contributions

Conceptualization: W.P.C., W.M.H.

Methodology: W.P.C., W.M.H., H.H.N., Y.Y.L., W.C.T., J.M.J.

Chemical synthesis: C.M.S.

Computer simulations of the protein structure: S.R.T.

Investigation: Y.Y.L., W.C.T., S.N.C., H.L.K., Y.S.S., Y.J.K., Y.Y.W., K.Y.K., Y.T.W., A.D.L., K.Y.K., Y.W.W.

Data analysis: L.C.L., M.H.L., Y.Y.L., W.P.C., W.C.T., H.L.K., R.H.

Clinical examination & echocardiography: W.C.T., S.N.C., M.H.W.

Funding acquisition: W.P.C., W.M.H., H.H.N.

Project administration: Y.Y.L., W.C.T.

Writing – original draft: Y.Y.L., W.C.T., W.P.C.

Writing – review & editing: W.P.C., W.M.H., H.H.N.

---

**Schematic diagram.**
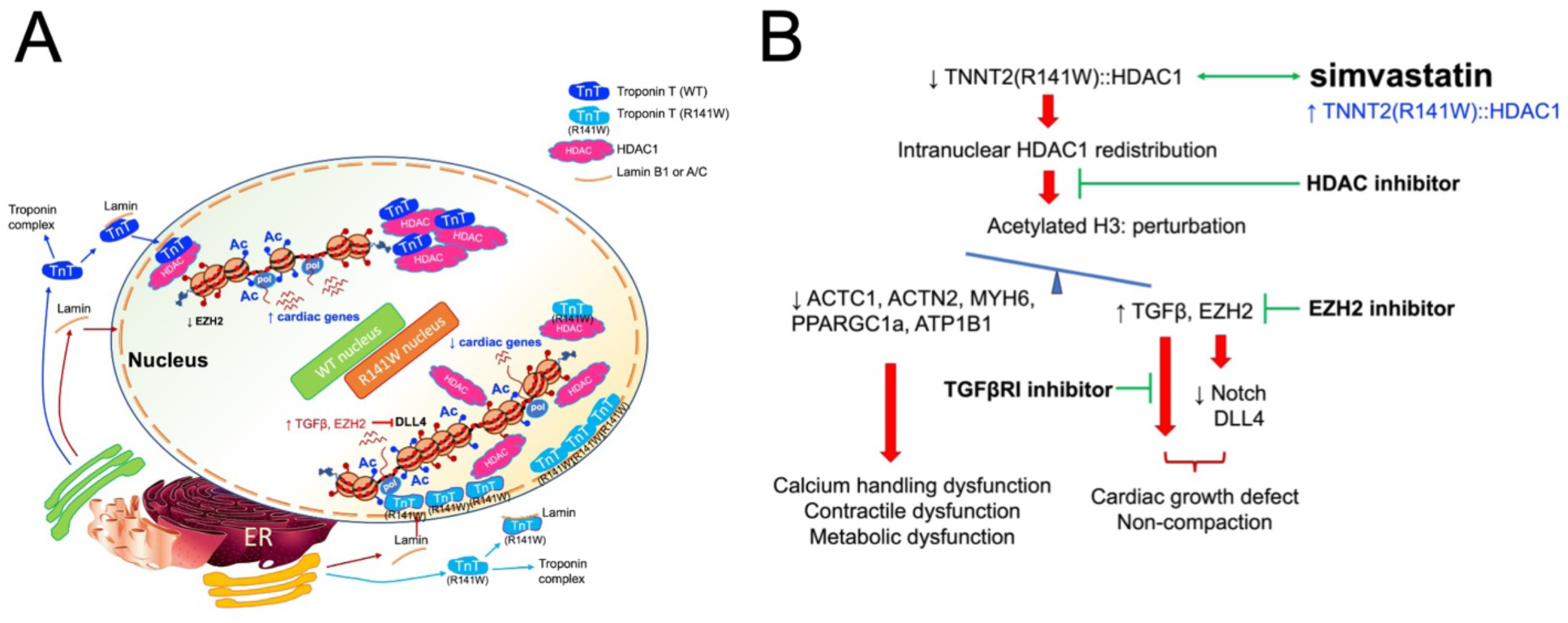
TNNT2(R141W)-mediated pathological signaling pathways and druggable targets. (A) Troponin T (TNNT2) can act as an HDAC1 sponge in the nuclei of healthy cardiomyocytes to maintain cardiac epigenetic and gene expressions. In contrast, a weakened association between mutant troponin T (TNNT2(R141W)) and HDAC1 within the nuclei of LVNC-cardiomyocytes could lead to the redistribution of nuclear HDAC1 to perturbate cardiac epigenetic. The decline in H3K27Ac marks within cardiac genes encoding the proteins functioning in muscle contraction and metabolism coincided with the downregulation of these affected genes and cardiac dysfunction. Concurrently, the increase in H3K27Ac marks within the promoter regions of TGF-β signaling molecules and EZH2 aligned with an abnormal upsurge in TGF-β stimulation and EZH2 upregulation. The increased EZH2 caused the downregulation of DLL4 expression. The excessive TGF-β stimulation and the diminished Notch DLL4 activity contribute to the decrease of myocyte proliferation and cardiac growth defect. (B) The administration of EZH2 inhibitor and TGF-βR1 inhibitor exhibited improvements in cardiac growth, while the use of an HDAC inhibitor partially ameliorated cardiac βAR responsiveness via inhibiting the redistributed HDAC1 to recover cardiac gene expression. Simvastatin demonstrated comprehensive improvement of cardiac growth and function through the restoration of the intranuclear TNNT2(R141W)-HDAC1 association.

