## Supplementary Figures & Tables for "Targeting mutant TNNT2-induced epigenetic perturbation and pathogenic signaling in left ventricular non-compaction cardiomyopathy"

**Supplementary figures & figure legends.**

**Supplementary figure 1**

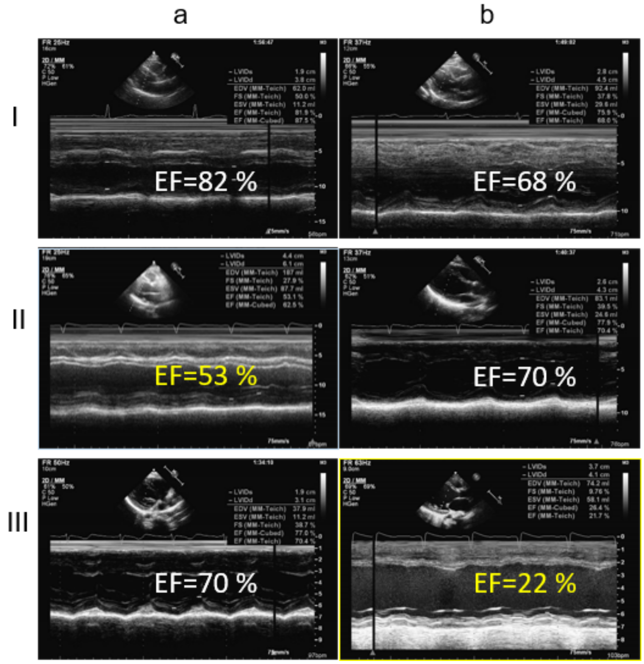

Supplementary Figure 1. The echocardiography of the LVNC family in 2015. Representative acquired images from the proband's grandfather (Ia, EF=82 %), grandmother (Ib, EF=68 %), father (IIa, EF=53 %), mother (IIb, EF=70 %), sister (IIIa, EF=70 %), and the proband (IIIb, EF=22 %).

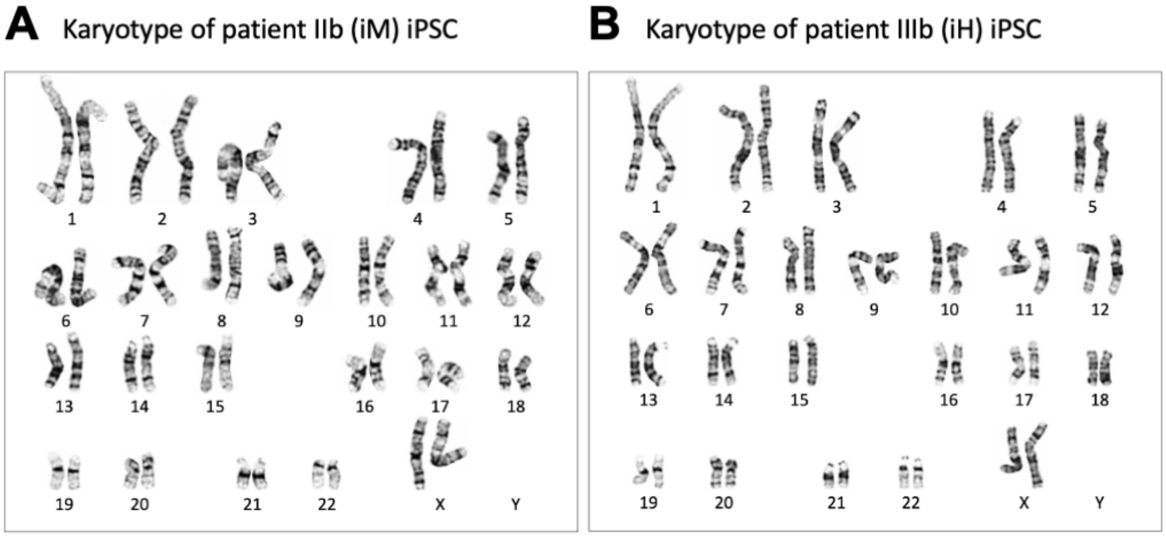

Supplementary Figure 2. Karyotype analysis of hiPSCs derived from (A) patient IIb and (B) patient IIIb demonstrated normal chromosomes in human iPSC lines.

### Supplementary figure 3

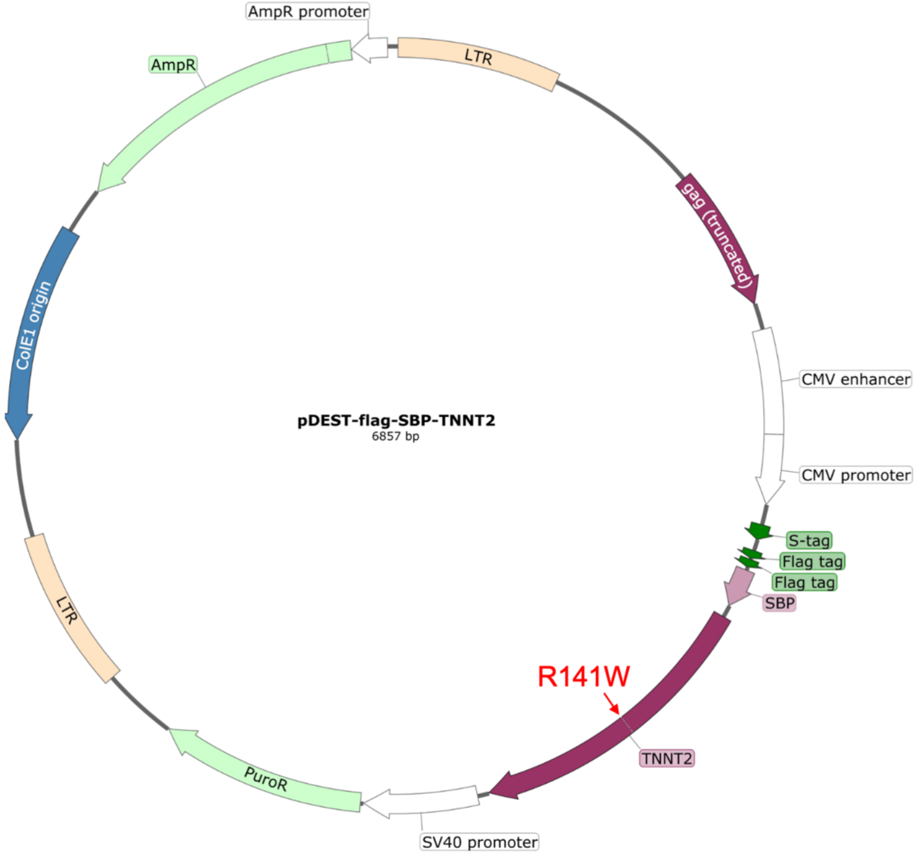

Supplementary Figure 3. The map of pDEST-*TNNT*2 construct coding the fusion proteins of flag-SBP-TNNT2(R141) or -TNNT2(W141) for pulldown assay of TNNT2 interaction proteins.

Supplementary Figure 4

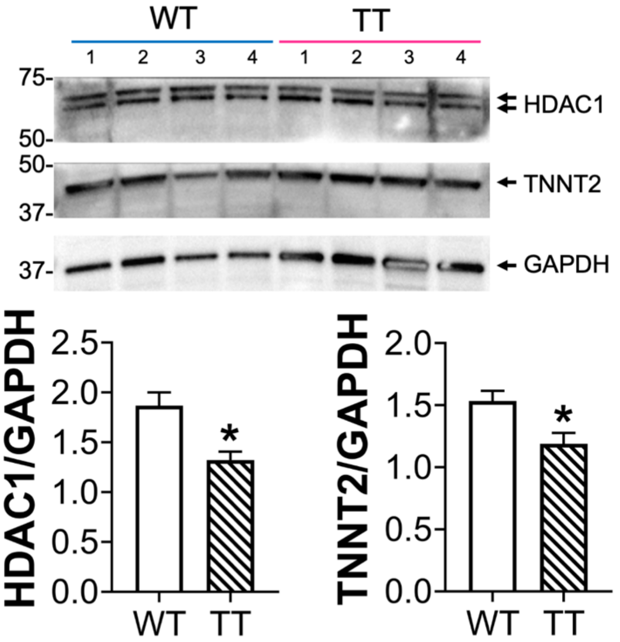

Supplementary Figure 4. Downregulation of HDAC1 and TNNT2 in TT (*Tnnt2*$^{R154W/R154W}$) mice hearts measured by western blot in comparison to those in WT (wild type) group. Data are mean±SEM (n=4 in each group). P<0.05, WT vs. TT by unpaired t test.

**Supplementary figure 5**

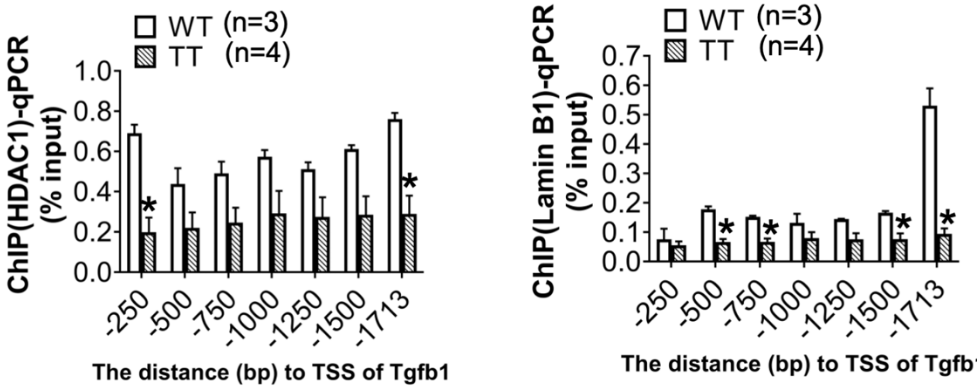

Supplementary Figure 5. Characterization of HDAC1 and lamin B1 marks in the promoter region of *Tgfβ*1 in WT (wild type) and TT (*Tnnt2*$^{R154W/R154W}$) mice hearts by ChIP-qPCR. Chromatin immunoprecipitation was performed using antibodies targeting HDAC1 or lamin B1, followed by qPCR quantification with normalization to the input. The data presented in this analysis represent the mean±SEM of three-four distinct mice hearts in each group. Statistical significance was assessed using an unpaired t test and values with *P<0.05 were considered significant when compared to the WT group at the same promoter region.

**Supplementary figure 6**

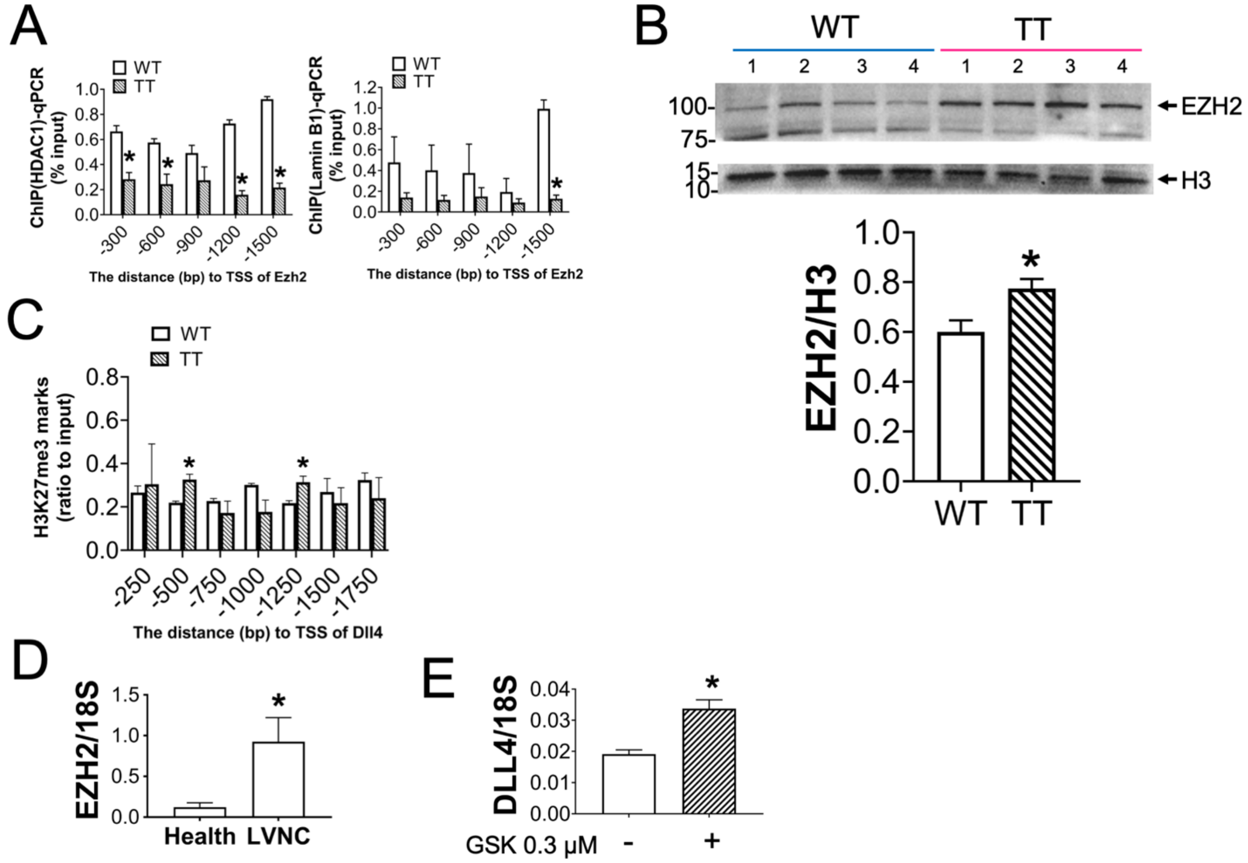

Supplementary Figure 6. (A) ChIP-qPCR was performed to explore less HDAC1 and lamin B1 marks within the promoter region (-1~-1500 bp from TSS) of *Ezh2*. (B) The upregulation of EZH2 measured by western blot in TT (*Tnnt2*$^{R154W/R154W}$) mice hearts in comparison to that in WT (wild type) group. Data are mean±SEM (n=4 in each group). P<0.05, WT vs. TT by unpaired t test. (C) The increase of H3K27me3 marks within the promoter region of DLL4 in TT mice hearts by ChIP-qPCR. Data are mean±SEM (n=3 in each group). *P<0.05 versus WT group by unpaired t test. (D) The upregulation of EZH2 in LVNC-hCM by absolute qPCR. Data are mean±SEM (n=4 in each group). *P<0.05, versus health-hCM by unpaired t test. (E) GSK503 (GSK, 0.3 μM) can increase DLL4 expression in LVNC-hCM by absolute qPCR. Data are mean±SEM (n=4 in each group). *P<0.05 versus control group by unpaired t test.

### Supplementary figure 7

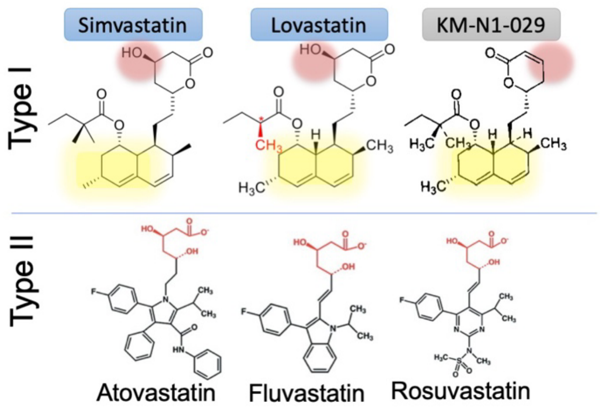

Supplementary Figure 7. Comparison of chemical structure differences among various statins. Notably, the structural motifs highlighted in vibrant yellow (naphthalene) and pink (mevalonolactone) hold pivotal significance in their capacity to enhance LVNC-hCM functionality.

### Supplementary Tables.

**Supplementary Table 1. Antibody list**

| REAGENT or RESOURCE | Host | SOURCE | IDENTIFIER |
|---|---|---|---|
| **Primary Antibodies** | | | |
| Beta actin [GT5512] | Mouse | GeneTex | GTX629630 |
| Cardiac Troponin T [1C11] | Mouse | GeneTex | GTX28295 |
| DDDDK tag | Rabbit | GeneTex | GTX115043 |
| DDDDK tag [FG4R] | Mouse | GeneTex | GTX82562 |
| EZH2 | Rabbit | GeneTex | GTX110384 |
| GAPDH | Rabbit | GeneTex | GTX100118 |
| HDAC1 | Rabbit | GeneTex | GTX100513 |
| Histone 3 | Rabbit | GeneTex | GTX122148 |
| Histone H3K27ac (Acetyl Lys27) | Rabbit | GeneTex | GTX128944 |
| Histone H3K27ac (Acetyl Lys27)- ChIP grade | Rabbit | GeneTex | GTX60815 |
| Histone H3K27me3 (tri-methyl Lys27) | Rabbit | GeneTex | GTX60892 |
| Lamin A/C | Rabbit | GeneTex | GTX101127 |
| Lamin B1 | Rabbit | GeneTex | GTX54288 |
| Cardiac Troponin T [1C11] | Mouse | GeneTex | GTX28295 |
| Cardiac sarcomeric actinin alpha | Mouse | GeneTex | GTX29465 |
| DLL4 [4A11] | Mouse | GeneTex | GTX60604 |
| DLL4 | Rabbit | Abcam | ab176876 |
| Jagged 1 | Rabbit | GeneTex | GTX48691 |
| Jagged 1 | Rabbit | Abcam | ab300561 |
| Recombinant Anti-HDAC1 + HDAC2 antibody [EPR20327] | Rabbit | Abcam | ab219054 |
| Histone H3(phosphor S10) | Rabbit | Abcam | Ab267372 |
| **Secondary Antibodies** | | | |
| HRP conjugated goat anti-mouse IgG | Goat | BioLegend | |
| HRP conjugated goat anti-rabbit IgG | Rabbit | BioLegend | |
| Alexa Fluor 488 goat anti-mouse IgG | Goat | BioLegend | |
| Alexa fluor 594 conjugated donkey anti-rabbit IgG | Donkey | BioLegend | |
| Alexa Fluor 647 donkey anti-rabbit IgG | Donkey | BioLegend | |

**Supplementary Table 2. Primers for ChIP-qPCR**

| Oligo ID | Sequence |
|---|---|
| F1-Tgfb1 prom1-250_97bp-mChIPqPCR | CCCACCCACTTACCTGCTGA |
| R1-Tgfb1 prom1-250_97bp-mChIPqPCR | CCAGGTCTGTAGTAGCCCTCA |
| F2-Tgfb1 prom251-500_142bp-mChIPqPCR | TTTGGGCTGGGTACTTGTGT |
| R2-Tgfb1 prom251-500_142bp-mChIPqPCR | CGGCCTGCACATACCTACTAC |
| F3-Tgfb1 prom501-750_137bp-mChIPqPCR | CCATCTTATCGGTCCAAGCCA |
| R3-Tgfb1 prom501-750_137bp-mChIPqPCR | GGAACCCTTCCTTCACCCG |
| F4-Tgfb1 prom751-1k_93bp-mChIPqPCR | TGTACTTAGGCCTGGTCCCTT |
| R4-Tgfb1 prom751-1k_93bp-mChIPqPCR | TCAGAACCTGGGAAATTTGGT |
| F5-Tgfb1 prom1k-1250_149bp-mChIPqPCR | GCCATCTGTCCATACCCATCT |
| R5-Tgfb1 prom1k-1250_149bp-mChIPqPCR | AGCCCGTTCCAAAATAACCC |
| F6-Tgfb1 prom1251-1500_90bp-mChIPqPCR | TATGTATGCCTCCTGCCGTT |

| | |
|---|---|
| R6-Tgfb1 prom1251-1500_90bp-mChIPqPCR | GCGGAATTTGGTTAGAGGCA |
| F7-Tgfb1 prom1501-1713_94bp-mChIPqPCR | CCACTCTGACTTGGTTTCAGC |
| R7-Tgfb1 prom1501-1713_94bp-mChIPqPCR | CAATCCAGCTTTGTGGCTCT |
| F1-Dll4prom1-250_117bp-mChIPqPCR | ACGCTCACAACCTCATGTTTCTTA |
| R1-Dll4 prom1-250_117bp-mChIPqPCR | CGTGGTAGGGGGAAGTGGAA |
| F2-Dll4 prom251-500_150bp-mChIPqPCR | CCACCGCTCTCACTGTAGG |
| R2-Dll4 prom251-500_150bp-mChIPqPCR | GCTGGCTTCTCCCAGTTTT |
| F3-Dll4 prom501-750_116bp-mChIPqPCR | CACGACGGGTTCTGGGAAAG |
| R3-Dll4 prom501-750_116bp-mChIPqPCR | CTGGAGGCGCATAGAGTTGT |
| F4-Dll4 prom751-1k_119bp-mChIPqPCR | CAGGTGAGATAGGCACTGAGG |
| R4-Dll4 prom751-1k_119bp-mChIPqPCR | AGAGCCAGGACTAGAGGTTCG |
| F5-Dll4 prom1k-1250_141bp-mChIPqPCR | AAGGTTAAGGCGTCGGATGG |
| R5-Dll4 prom1k-1250_141bp-mChIPqPCR | AAAAAGCAGGACCAGGGGAT |
| F6-Dll4 prom 1251-1k5_104bp-mChIPqPCR | ATTCGGGAACCACCTCACCT |
| R6-Dll4 prom 1251-1k5_104bp-mChIPqPCR | GCGAGCAGTCCTTAGAGATCG |
| F7-Dll4 prom 1501-1750_135bp-mChIPqPCR | ACTGACAGGCTGCGAAGAG |
| R7-Dll4 prom 1501-1750_135bp-mChIPqPCR | AGTACACTGCACCCAAGTGAC |
| F1-Gapdh prom1-250 bp_111bp-mChIPqPCR | TGCCTCATCCTTGCACAACT |
| R1-Gapdh prom1-250 bp_111bp-mChIPqPCR | CCGAATGTTTTCTGGGGTGC |
| F2-Gapdh prom251-500 bp_105bp-mChIPqPCR | CCCGAAGACTGGCAAATGACT |
| R2-Gapdh prom251-500 bp_105bp-mChIPqPCR | TAGTGAAGGACTGGCTCTGGG |
| F3-Gapdh prom501-750 bp_104bp-mChIPqPCR | GTGCTCCCTACCTCTTGGTG |
| R3-Gapdh prom501-750 bp_104bp-mChIPqPCR | TTCCATAGCCCAGCCCTAAC |
| F4-Gapdh prom751-1Kbp_102bp-mChIPqPCR | TCGGTAGGAAGGCAGAAAAGG |
| R4-Gapdh prom751-1Kbp_102bp-mChIPqPCR | CAGGCCAGATGAACAGGGTG |
| F5-Gapdh prom1K-1250bp_123bp-mChIPqPCR | CATGTTGCACTGGCCTAGCA |
| R5-Gapdh prom1K-1250bp_123bp-mChIPqPCR | CGAGACCGGGATTCTTCACTC |
| F6-Gapdh prom1251-1553bp_99bp-mChIPqPCR | TCTCCTGTGTTCTCCCCTCAC |
| R6-Gapdh prom1251-1553bp_99bp-mChIPqPCR | ATCCAGGGACGTGCTGACTG |
| F1-mEzh2-prom 0-300_94bp | GTTCGGCCCTCTGATTGG |
| R1-mEzh2-prom 0-300_94bp | ATCGCCATCGCTTTTATTTG |
| F2-mEzh2-prom -301_600_99bp | TCACACGCCTTCCTTTCAGT |
| R2-mEzh2-prom -301_600_99bp | TCGGGTTGGTAACGGTCTTA |
| F3-mEzh2-prom -601_900_90bp | ACATGGGGTGAGCTATTTGC |
| R3-mEzh2-prom -601_900_90bp | CCTTCCAAGCTGCGTTTATG |
| F4-mEzh2-prom -901_1k2_100bp | ACAGGTCTGTGGGATTTTGC |

| | |
|---|---|
| R4-mEzh2-prom -901_1k2_100bp | CCCCACTCCCTGTTCAGTTA |
| F5-mEzh2-prom -1k2-1k5_100bp | TTACCAACGGAAAACCCAAG |
| R5-mEzh2-prom -1k2-1k5_100bp | TCACTGGCATTAGAGCACACA |

**Supplementary Table 3. Primers for qPCR**

| Oligo ID | Sequence | Source |
|---|---|---|
| Human qPCR primers | | |
| F-hDLL4-117bp | CTGTG CAAGA AGCGC AATGA | This study |
| R-hDLL4-117bp | GCCCG AAAGA CAGAT AGGCT G | This study |
| F1-h-18srRNA-123bp | GCAATTATTCCCCATGAACG | This study |
| R1-h-18srRNA-123bp | GGCCTCACTAAACCATCCAA | This study |
| F01-hEZH2-147bp-exon4_5 | TTCTTGGTCTCCCCTACAGC | This study |
| R01-hEZH2-147bp-exon4_5 | TCCCCGTGTACTTTCCCATC | This study |
